# Inferring rheumatoid arthritis disease activity status from the electronic health records across health systems to enable real-world data studies

**DOI:** 10.1101/2025.11.13.25340003

**Authors:** David Cheng, Xuan Wang, Gregory C. McDermott, Jennifer S. Hanberg, Zoe Love, Katherine Zhong, Mary Jeffway, Jue Hou, Vidul Panickan, Rahul Sangar, Ying Qi, Connor Melley, Lauren Costa, Dakota Feil, Rachael Matty, Dana Weisenfeld, Abisayo Animashaun, Aimee Schreiner, Sara Morini, Lauren Rusnak, Andrew Cagan, Misti Paudel, J. Michael Gaziano, Brian Sauer, Michael Weinblatt, Joshua Baker, Bryant England, Yuk-Lam Ho, Kelly Cho, Paul Monach, Grant W. Cannon, Nancy Shadick, Ted R. Mikuls, Tianxi Cai, Katherine P. Liao

**Author notes:** Correspondence: Katherine P. Liao, MD, MPH VA Boston Healthcare System 150 South Huntington Ave Jamaica Plain, MA 02130. contributed equally (David Cheng, Xuan Wang, Tianxi Cai, Katherine P. Liao).

## Abstract

**Objective:** Disease activity plays a central role in rheumatoid arthritis (RA) clinical studies. However, RA disease activity is inconsistently recorded in real-world electronic health records (EHR) data limiting the generation of real-world evidence (RWE). This study aimed to develop and validate scalable machine learning (ML) models to infer RA disease activity from EHR data.

**Methods:** We conducted studies from EHR data from Mass General Brigham (MGB) and the Veterans Affairs (VA); both have RA registries with prospectively collected disease activity score 28 (DAS28). The features for the algorithm were extracted from the EHR including structured data, e.g., ICD codes and narrative data using natural language processing (NLP). Machine learning models were trained on the registry-collected DAS28.We tested within-institution trained model performance and across systems transportability. The association between inferred disease activity and major adverse cardiovascular events (MACE) was tested with stratified Cox models to test face-validity.

**Results:** We studied 1105 MGB and 2631 VA RA patients. Models with structured data models achieved an AUC of 0.68-0.70; models incorporating structured and NLP achieved higher performance (AUC=0.843, MGB; 0.833, VA). Cross-site validation demonstrated reduced transportability (AUC=0.679, MGB→VA; 0.718, VA→MGB), due to differences in the important feature. Within institution, inferred disease activity was significantly associated with increased risk for incident MACE (MGB: HR=1.12; VA: HR=1.14).

**Conclusion:** RA disease activity can be inferred at scale from within-institution EHR data, though cross-institution performance is limited. The inferred disease activity replicated association between RA and MACE and supports it’s use in future studies to generate RWE.

## Introduction

Measurement of disease activity in patients with rheumatoid arthritis (RA) is a key variable in clinical and epidemiologic research and is recommended as part of clinical management [1]. RA disease activity is measured by several validated indices such as the Disease Activity Score with 28-joint counts based on C-reactive protein (DAS28-CRP)[2]. In studies relying upon electronic health records (EHR) data, measures of disease activity are often not directly observed, although EHRs capture data on patient symptoms, physical examinations, labs, and medications that correlate with disease activity. The lack of systematically collected disease activity data to use as an exposure, outcome, or confounder is a major barrier in leveraging large-scale real-world data (RWD) from EHRs to generate real-world evidence in RA. For example, RA disease activity is frequently used as an outcome in drug trials, including those used for regulatory approvals in the United States and Europe. Studies have demonstrated associations between higher RA disease activity as an exposure with safety and effectiveness outcomes such as major adverse cardiovascular events (MACE) [3,4]. RA treatment guidelines have also suggested treat-to-target approaches whereby clinicians are encouraged to formally assess disease activity measures and intensify therapy until achieving low disease activity or remission [1,5].

Models for predicting disease activity using EHR data in specific settings have been previously attempted. An early study using natural language processing (NLP) leveraged concepts identified from clinical notes along with lab values to infer disease activity at rheumatology clinic visits [1,6]. Later work has demonstrated that models based on concepts from notes alone can achieve good performance when trained on a large sample based on registry data [8]. Another study linking claims to EHR data has found that the addition of lab data from EHRs significantly improved the performance of models for disease activity [9]. The ability to forecast future disease activity using structured data at a given clinical visit, including demographics, medications, labs and prior disease activity measures has also been reported [10]. Other recent works have developed models that incorporated patient-reported outcomes [11] and specifically for patients treated with biologics [12]. Despite these existing models, there is a lack of recent algorithms for EHR data that incorporate both structured and narrative data and allow for disease activity to be inferred at specific times during patient follow-up, which would enable use of disease activity as either an outcome, exposure, or covariate in studies with longitudinal follow-up.

In this study, we focus on retrospectively inferring disease activity status using both structured and narrative data from clinical notes in EHRs from the Mass General Brigham (MGB) system and the Department of Veterans Affairs (VA) [13,14]. Both MGB and the VA have RA registries linked to EHRs: the Brigham Rheumatoid Arthritis Sequential Study (BRASS) and VA Rheumatoid Arthritis Registry (VARA). Both registries prospectively collected DAS28-CRP. The linkage between the registry and EHR enables the creation of a platform to train algorithms to predict disease activity collected in registries using EHR data.

The objective of this study was to develop models that allow for time-specific inferences over specified months during patients’ EHR follow-up. We tested the feasibility of this approach at MGB and the VA, training models for each institution and testing the transportability of the models both within and across institutions. We hypothesized that the addition of NLP to structured EHR data would significantly improve the accuracy for inferring disease activity. Additionally, we sought to replicate the known association between disease activity and future MACE as an example of a downstream application using inferred disease activity [15,16].

## Patients and Methods

### Patients

We identified patients in the EHRs at each institution with an ICD-9/10 code for RA (ICD-9: 714.X and ICD-10: M06.9 and local codes) who were participants of the corresponding registry study at each site. Individuals enrolled in the Brigham and Women’s Rheumatoid Arthritis Sequential Study (BRASS) [17] with prospectively collected data from 2003-2019 were linked with data from the MGB EHR. For VA, we studied individuals enrolled in the Veteran Affairs Rheumatoid Arthritis registry (VARA) using data from 1999-2020 [18] with data linked to the VA EHR. Patients with ≥1 measurement of DAS28-CRP at a registry visit were included in the study population for the analysis [19]. The relationships between the registries, EHR and an overview of the study design are outlined in **Figure 1a**. In the downstream analysis assessing associations of inferred disease activity with subsequent MACE, we identified a broader RA population from the EHRs using a validated multimodal RA phenotyping algorithm [20] that was separately applied at both institutions. RA patients in that analysis were identified as those with a predicted probability for having RA above a threshold such that the positive predictive value (PPV) is ≥90% at each site.

**Figure 1.**
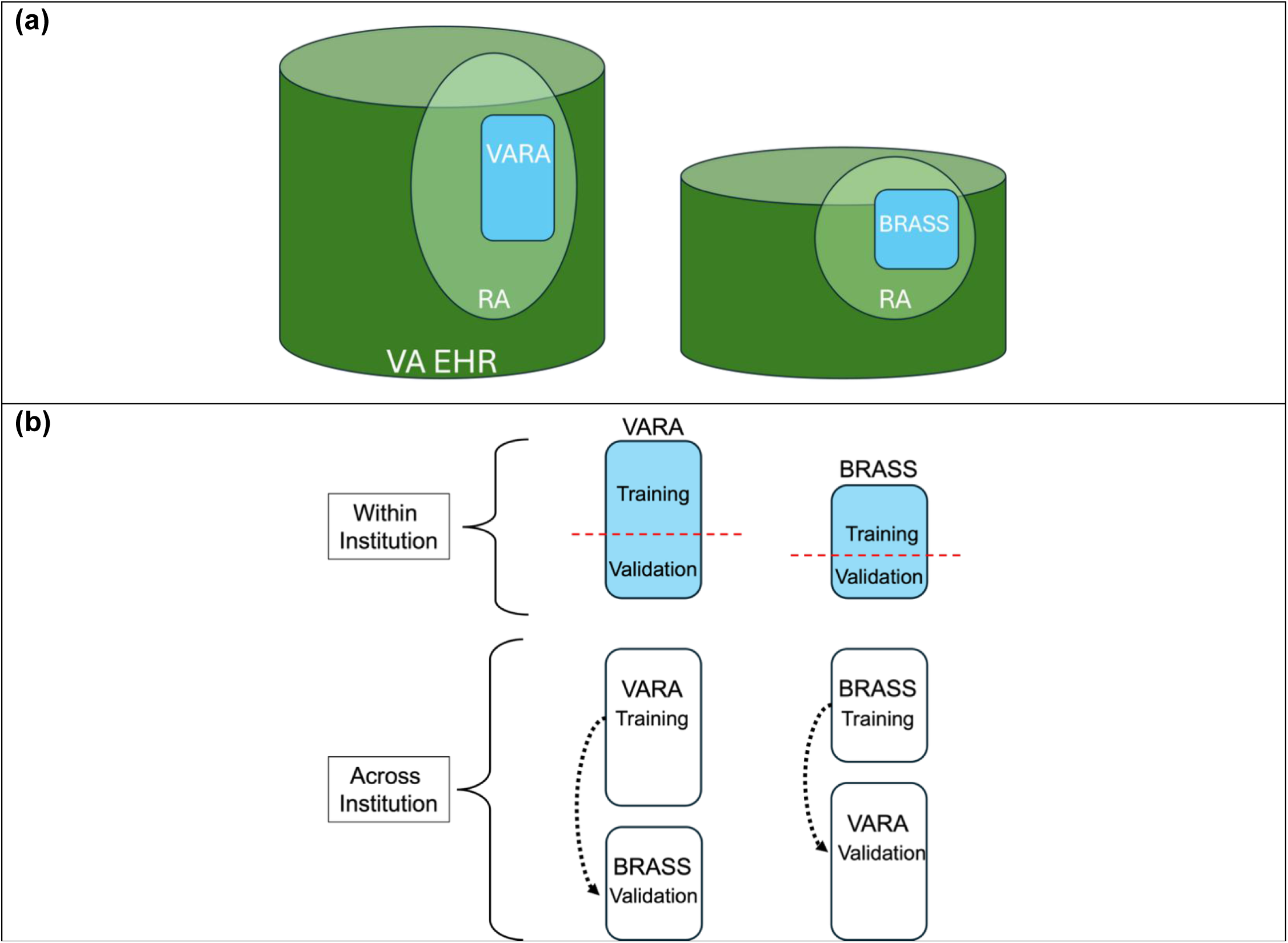
(a) Overview of relationships between the institution specific registries, BRASS and VARA, which contain DAS28-CRP data that are consistently collected through research visits, electronic health record (EHR) based RA cohorts identified using a phenotyping pipeline (light green circle), and the overall EHR data; and **(b)** the study design to develop models for disease activity within and across institutions.

### Observed Disease Activity Status

The DAS28-CRP was prospectively assessed at registry visits in BRASS and VARA. We identified months in which DAS28-CRP was assessed for patients in the study sample and dichotomized DAS28-CRP at 3.2 to classify patients to moderate or high disease activity (DAS28-CRP >3.2) versus remission or low disease activity (DAS28-CRP ≤3.2) in the month. When multiple assessments were recorded within a month, we took the median of available measurements and then dichotomized to define disease activity status in the month.

### EHR Features

We extracted data on patients’ demographics and structured data, i.e., codes related to RA including diagnosis, procedure, medication, and laboratory codes. These codes were obtained from a knowledge network that identifies codes related to phenotypes in the EHR [21–24]. Additionally, available data on 4 laboratory values considered important in assessing RA and disease activity were extracted: C-reactive protein (CRP), erythrocyte sedimentation rate (ESR), antibodies to cyclic-citrullinated peptide (anti-CCP), and rheumatoid factor (RF). All available laboratory data were initially extracted for all patients on all dates. For ESR and CRP, the data were aggregated into monthly counts of all available codified data based on observed dates. Laboratory values were then summarized based on the median values in months with a measurement. Data harmonization was required for CRP which were reported in both mg/dL and mg/L. RF and anti-CCP status were defined as positive if an individual had a positive test in the month.

All clinical notes were processed using natural language processing (NLP) to identify mentions of RA-and disease activity-related concepts from the narrative clinical notes. A dictionary of RA-related concepts was created through the Online Narrative and Codified feature Search Engine (ONCE) tool generated by the knowledge network mentioned above [20]. A separate NLP dictionary was developed to identify terms associated with disease activity. We performed a manual review of rheumatology notes and sections of text that were informative for determining disease activity were flagged. Named entity recognition was applied to identify clinical terms from these text. The terms were mapped to NLP concepts and associated concept unique identifiers (CUI) were identified using the Unified Medical Language System. All concepts not already present in the RA dictionary generated by ONCE were retained in the disease activity NLP dictionary.

The monthly count of mentions of concepts from both the RA and disease activity dictionaries were then obtained based on the date of the notes. We refer to the counts of codes as *codified features*, the codified and lab value data as *structured features*, and the counts of NLP concepts as the *narrative features*. A comprehensive list of the structured features can be found in **Supplemental Table 1**, and NLP features in **Supplemental Table 2.**

### Modeling approach for inferring RA disease activity

The approach for inferring disease activity is illustrated in **Figure 2a**. For each month in which DAS28-CRP was measured in the registry (target month), we further summarized the EHR features that are dated to be within 3 months (i.e., ≥ (target month - 3 months) or ≤ (target month + 3 months)). The codified and narrative features were aggregated into total counts of specific codes and CUIs incurred within this time window, respectively. The CRP and ESR data were further summarized by the median value over the window, and the anti-CCP and RF were summarized by presence or absence of positive test over the window. Missing CRP and ESR values were imputed by single imputation using a LASSO model given other screened features (see the following paragraph) in target months with no observed lab value. All these features were then used to train a model to predict the disease activity status in the target months.

**Figure 2.**
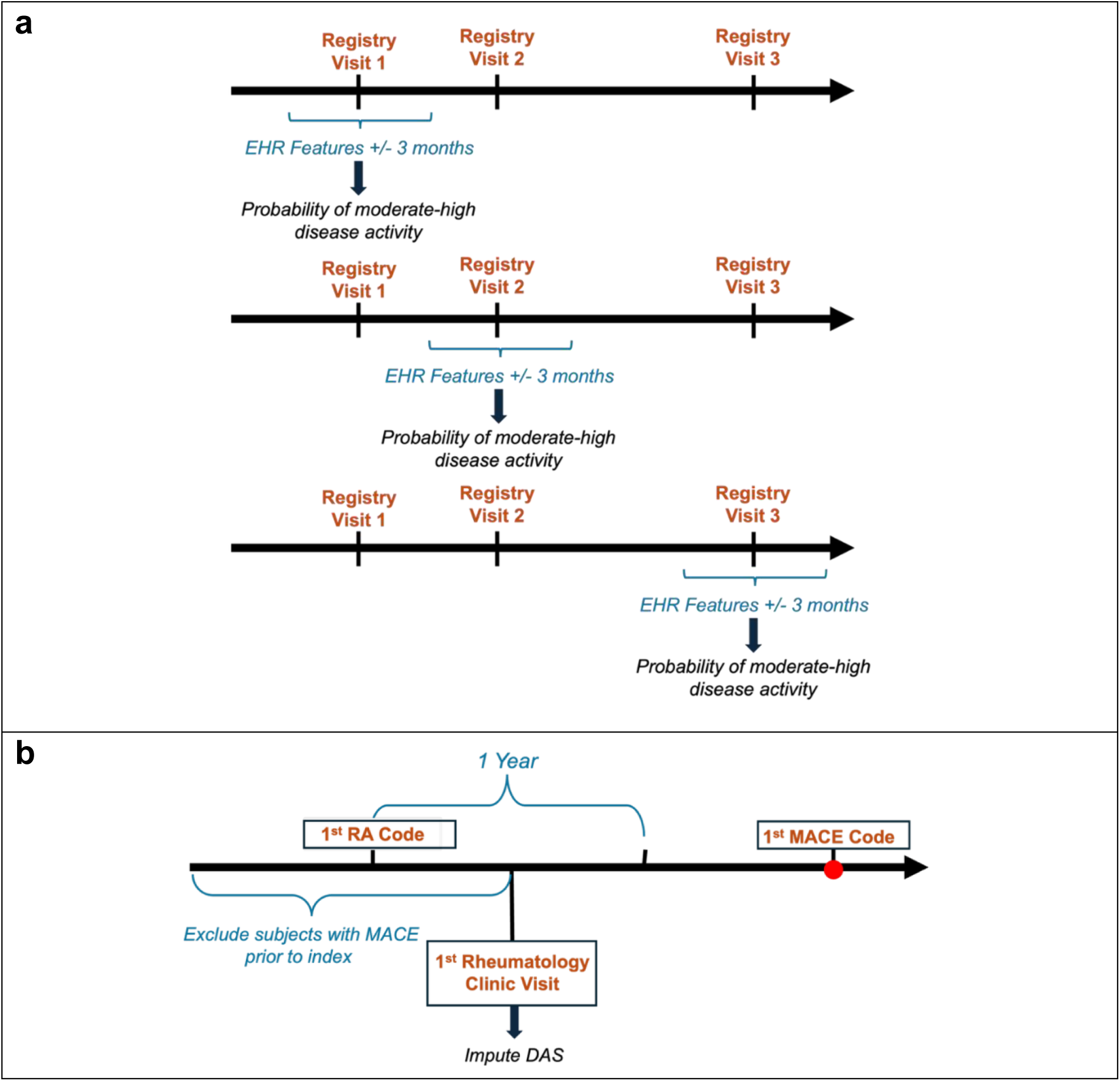
(a) Approach for training time-specific disease activity inferences over specified months during EHR follow-up, and **(b) s**tudy design for applying inferred RA disease activity to replicate the association between disease activity and MACE in RA. The 1^st^ RA code refers to the first RA diagnosis code observed in EHR follow-up. Patients with at least one rheumatology clinic visit within 1 year after the 1^st^ RA code are included in the analysis. Those with any MACE events observed prior to the 1^st^ rheumatology clinic visit during EHR follow-up were excluded.

To identify features that are potentially relevant to disease activity based on the existing data, feature screening was performed. This was done by fitting a multivariable logistic regression model for moderate-high disease activity at target months against each individual candidate feature while additionally adjusting by age, sex, race, and healthcare utilization, where healthcare utilization is defined as the number of days with any ICD code [25] recorded during EHR follow-up. Codified and narrative features with p-values ≤0.1 and prevalence >5% were then selected to be features used for training the disease activity algorithm (**Supplemental Figures 1-4**). We used target months before

2009 for feature screening and reserved the data from 2009 onward for training and validation. This approach allowed us to separately leverage historical data for screening and 10 years of contemporary data for training and validation to avoid overfitting.

We fitted an ensemble of machine learning models using Super Learner [26] to estimate the probability of having moderate-high disease activity status (DAS28-CRP > 3.2) at each target month given the EHR features: the demographic (age, sex, race), healthcare utilization, laboratory value (ESR, CRP, RF and anti-CCP), and screened codified and narrative features. The ensemble included the mean, random forest [27], XGBoost [28], and neural network [29]. A weighted average of the estimated probabilities based on each component of the ensemble is taken to obtain the final probability estimates. These estimated probabilities measure how likely a patient is to have moderate or high disease activity. The probabilities can then be dichotomized at different thresholds to yield classification of patients into moderate-high versus remission-low disease activity status. When averaged across a sample of patients, the mean predicted probabilities represent an estimate of the prevalence of moderate-high disease activity status in the corresponding population.

### Evaluation of RA disease activity phenotyping model

We evaluated the phenotyping model when training and validating using data from the same institution. We also evaluated the performance when training in one institution and externally validating in another institution (**Figure 1b**). For within-institution evaluations, we randomly sampled 80% of the target months for training the phenotyping model and used the remaining 20% as an independent validation sample. All screened features available at each site were considered as candidate features for training and validation. For cross-institution evaluations, we trained the model using all target months from one institution and validated in target months from the other institution, restricting the set of screened features to those common to both sites.

To assess the incremental impact of different sets of features on the phenotyping performance in terms of discrimination, we evaluated the area under the receiver operating curve (AUC) in the validation sample for 5 models trained on: (1) codified features only, (2) codified features plus lab values, (3) codified data plus narrative features, (4) codified features plus both lab values and narrative features, and (5) codified features plus both lab values and narrative features that are common to both sites. We additionally assessed the phenotyping performance in terms of calibration by plotting the observed proportion with moderate-high disease activity (i.e., the prevalence of moderate-high disease activity) for patients within each decile of the predicted probability of moderate-high disease activity. For both the within- and cross-institution evaluations, we assessed the AUC and calibration plots in the corresponding validation samples. The relative importance of features for the algorithm was ranked using the SHapley Additive exPlanations (SHAP) values, which was calculated in terms of each feature’s contribution to reducing Mean Absolute Error (MAE, **Supplemental Figures 1-4**). The SHAP value provides a way to attribute the contribution of each feature toward the prediction outcome of a model [30].

### Application of inferred disease activity in an association study with major adverse cardiovascular events (MACE)

To demonstrate the application of the disease activity algorithm in a downstream analysis and assess its face validity, we tested the known association between higher disease activity with the risk of major adverse cardiovascular events (MACE) in RA [31]. We conducted this analysis in both MGB and VA data and restricted the populations to patients with at least one rheumatology clinic visit within 1 year after the first available RA diagnosis code (**Figure 2b**). Patients with diagnosis codes for MACE at any time prior to the first rheumatology clinic visit were excluded. We applied the model trained using target months linked to the respective registry at each institution to the broader cohort of

RA patients in the EHR data in the same institution (**Figure 1a**). The predicted probability of moderate-high disease activity was inferred for the month of the first rheumatology clinic encounter within 1 year after the first RA diagnosis code (index date).

The inferred moderate-high vs remission-low disease activity status was defined by dichotomizing the predicted probabilities at thresholds of 0.35 for MGB and 0.36 for VA, to match the prevalences of moderate-high disease activity status observed in BRASS and VARA. MACE events were ascertained based on the presence of diagnosis and procedure codes for myocardial infarction, ischemic stroke, use of coronary artery bypass graft, percutaneous coronary intervention, percutaneous transluminal coronary angioplasty, or stents [32] (**Supplemental Table 3**). Time-to-MACE was censored at the month of either the last structured or narrative feature. Kaplan-Meier analysis was performed to estimate the unadjusted MACE-free survival among those with inferred moderate-high vs remission low disease activity status. A stratified Cox model was fit to further assess this association after adjustment for patients’ age, sex, and self-reported race and stratifying the baseline hazard by calendar year of the rheumatology clinic visit.

## Results

We identified 1,105 participants in BRASS who were followed in the MGB system and had at least one measurement of DAS28-CRP in 5,072 distinct post-2009 target months (**Table 1**). The mean age in the month of the first post-2009 BRASS target month was 57.7 years, 82.4% of patients were female, and 87.1% were White; 85.7% were seropositive. Among VARA patients, 2,631 patients had a measurement of DAS28-CRP in 31,440 distinct post-2009 target months. The mean age in the month of the first post-2009 VARA visit was 64.6 years, 12.1% of patients were female, and 76.8% were White; 88.6% were seropositive. The proportion of target months in which patients were in remission, low, moderate and high disease activity were similar in BRASS and VARA. In the majority of target months, 59.8% of target months in BRASS and 60.6% in VARA, patients were in remission. There were 22.0% of target months in BRASS and 22.9% in VARA in which patients had moderate disease activity.

**Table 1.** Characteristics at the baseline visit for BRASS or VARA for the phenotyping analysis, and for the baseline for the MACE analysis.

|  | Disease activity phenotyping |  | MACE analysis |  |
| --- | --- | --- | --- | --- |
|  | MGB BRASS | VA VARA | MGB EHR with RA | VA EHR with RA |
| ***Patient-level*** |  |  |  |  |
| Patients, n | 1,105 | 2,631 | 16,553 | 72,847 |
| Age in first visit month, mean (SD) | 57.74 (13.48) | 64.57 (10.81) | 60.58 (17.85) | 61.90 (12.35) |
| Female, n (%) | 911 (82.44) | 318 (12.09) | 12995 (78.51) | 9757 (13.39) |
| Race/ethnicity, n (%) |  |  |  |  |
| White | 963 (87.14) | 2021 (76.81) | 12949 (78.23) | 51503 (70.70) |
| Black | 44 (3.98) | 436 (16.57) | 1004 (6.07) | 10813 (14.84) |
| Others | 30 (2.71) | 43 (1.63) | 691 (4.17) | 1735 (2.38) |
| NULL | 68 (6.15) | 131 (4.98) | 1909 (11.53) | 8796 (12.07) |
| Seropositivity status, n (%) | 937/1093 (85.73) | 1866/2107 (88.56) | 9031/13110 (68.89) | 7313/9976 (73.31) |
| ***Patient-visit/month-level*** |  |  |  |  |
| Target months with DAS28-CRP measured, n | 5,072 | 31,440 | 16,553 | 72,847 |
| Target months since first RA code at visit, mean (SD) | 133.77 (71.20) | 121.55 (58.58) | 1.56 (1.73) | 2.26 (2.39) |
| Concomitant medications at visit, n (%) |  |  |  |  |
| bDMARD | 1252 (24.68) | 9141 (29.08) | 2157 (13.03) | 7174 (9.85) |
| csDMARD | 305 (6.01) | 15674 (49.85) | 7672 (46.35) | 48846 (67.05) |
| tsDMARD | 0 (0) | 0 (0) | 107 (0.65) | 207 (0.28) |
| DAS28-CRP at visit, mean (SD) | 2.58 (1.18) | 2.51 (1.23) |  |  |
| DAS28-CRP Category at visit, n (%) |  |  |  |  |
| High disease activity (> 5.1) | 202 (3.98) | 1222 (3.87) |  |  |
| Moderate disease activity (3.2 - 5.1) | 1116 (22.0) | 7191 (22.87) |  |  |
| Low disease activity (2.6 - 3.2) | 721 (14.22) | 3989 (12.69) |  |  |
| Remission (≤ 2.6) | 3033 (59.8) | 19038 (60.55) | | |

[1] BRASS and VARA cohorts refer to participants of each registry with visits in which DAS28-CRP was prospectively collected and had linked EHR data.
[2] RA EHR cohorts refer to a broader cohort of RA patients at each institution included for the MACE analysis.
[3] DAS28-CRP was not available among RA EHR patients.
[4] Seropositivity status based on any RF or CCP positive test observed during EHR follow-up.

When separately trained and validated within MGB and the VA, the phenotyping models exhibited AUCs of 0.676 at MGB and 0.702 at the VA when using codified data only (**Table 2**). The AUC improved to 0.756 at MGB and 0.736 at the VA when additionally incorporating lab values and to 0.821 at MGB and 0.832 at the VA when additionally incorporating narrative features. The best model performance was observed when all codified and narrative features were included with an AUC of 0.843 at MGB and 0.833 at the VA.

**Table 2.** Within- and cross-institution AUC (95% CI in bracket) by feature set.

| **Feature Set** | **MGB** | **VA** |
| --- | --- | --- |
| (1) Codified (diagnosis, procedure, lab encounter, medication) | 0.676<br>(0.639-0.713) | 0.702<br>(0.687-0.717) |
| (2) Codified + Lab values (CRP, ESR, RF-pos, CCP-pos) | 0.756<br>(0.724-0.789) | 0.736<br>(0.722-0.750) |
| (3) Codified + NLP | 0.821<br>(0.794-0.848) | 0.832<br>(0.821-0.843) |
| (4) Codified + NLP + Lab values | 0.843<br>(0.819-0.868) | 0.833<br>(0.823-0.844) |

**Cross-Institution AUC**
| **Feature Set** | **Train at MGB**<br>**Test at VA** | **Train at VA**<br>**Test at MGB** |
| --- | --- | --- |
| (5) Codified + NLP + Lab values (common features) | 0.679<br>(0.672-0.685) | 0.718<br>(0.702-0.733) |

In cross-institution evaluations, applying the model trained using features common to both MGB and VA to an external site, the performance exhibited considerable degradation. Even when including both narrative and lab features, the AUC was 0.679 when trained at MGB and validated at the VA; the AUC was 0.718 when trained at VA and validated at MGB. While CRP, ESR, age and prednisone use were among the top 5 most important features at both institutions, the order of importance differed at MGB vs the VA (**Supplemental Figures 1-4**).

The observed probabilities of moderate-high disease activity exhibited relatively close agreement with mean predicted probabilities when models were trained on data from the same institution (**Figure 3**). There was some systematic under-estimation of the probability of moderate-high activity when training the model at the VA and validating at MGB. Conversely, there was over-estimation of the probability of moderate-high disease activity in most deciles when training the model at MGB and validating at the VA.

**Figure 3.**
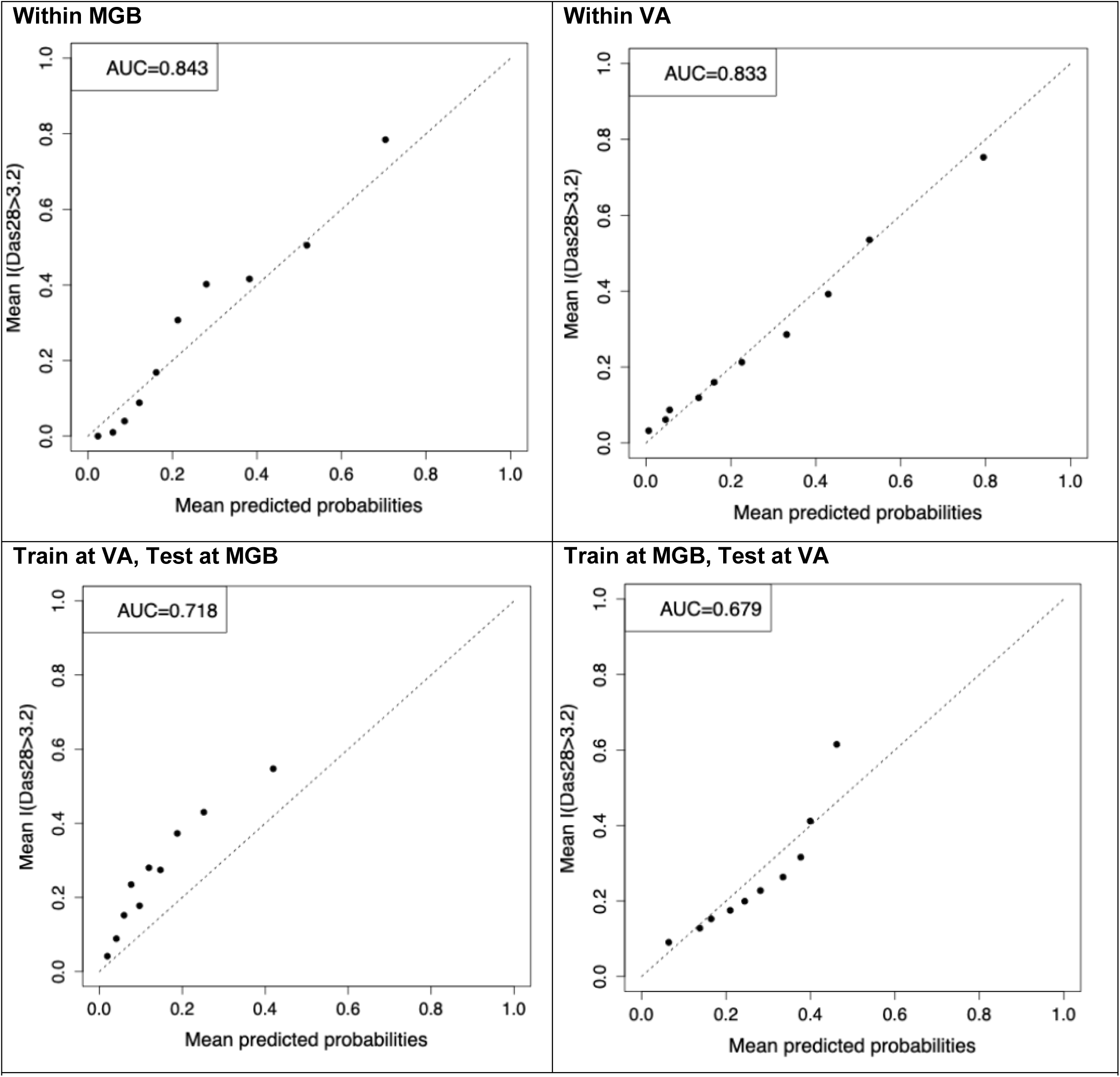
Within- and cross-institution calibration plots at MGB and VA.

In the analysis of associations between disease activity and MACE, we identified 16,553 patients at MGB and 72,847 at the VA with a rheumatology clinic visit within 1 year after the first RA diagnosis code without a prior MACE event. For the MGB population, at the index date, the mean age was 60.6 years, 78.5% were female, 68.9% seropositive, and 44.9% had an inferred moderate-high disease activity status with the chosen cutoff. For the VA population, at the index date, the mean age was 61.9 years, 13.4% were female, 73.3% seropositive, and 15.9% had an inferred moderate-high disease activity status with the chosen cutoff (**Table 1**). Patients with inferred moderate-high disease activity exhibited a higher risk for MACE at both MGB and VA (**Figure 4**). In Cox analyses adjusted for baseline covariates (age, gender, race) and stratified by calendar year, inferred moderate-high disease activity status was also significantly associated with higher hazard of subsequent MACE at both MGB (HR=1.12, 95% CI 1.00-1.25) and VA (HR=1.14, 1.08-1.21).

**Figure 4.**
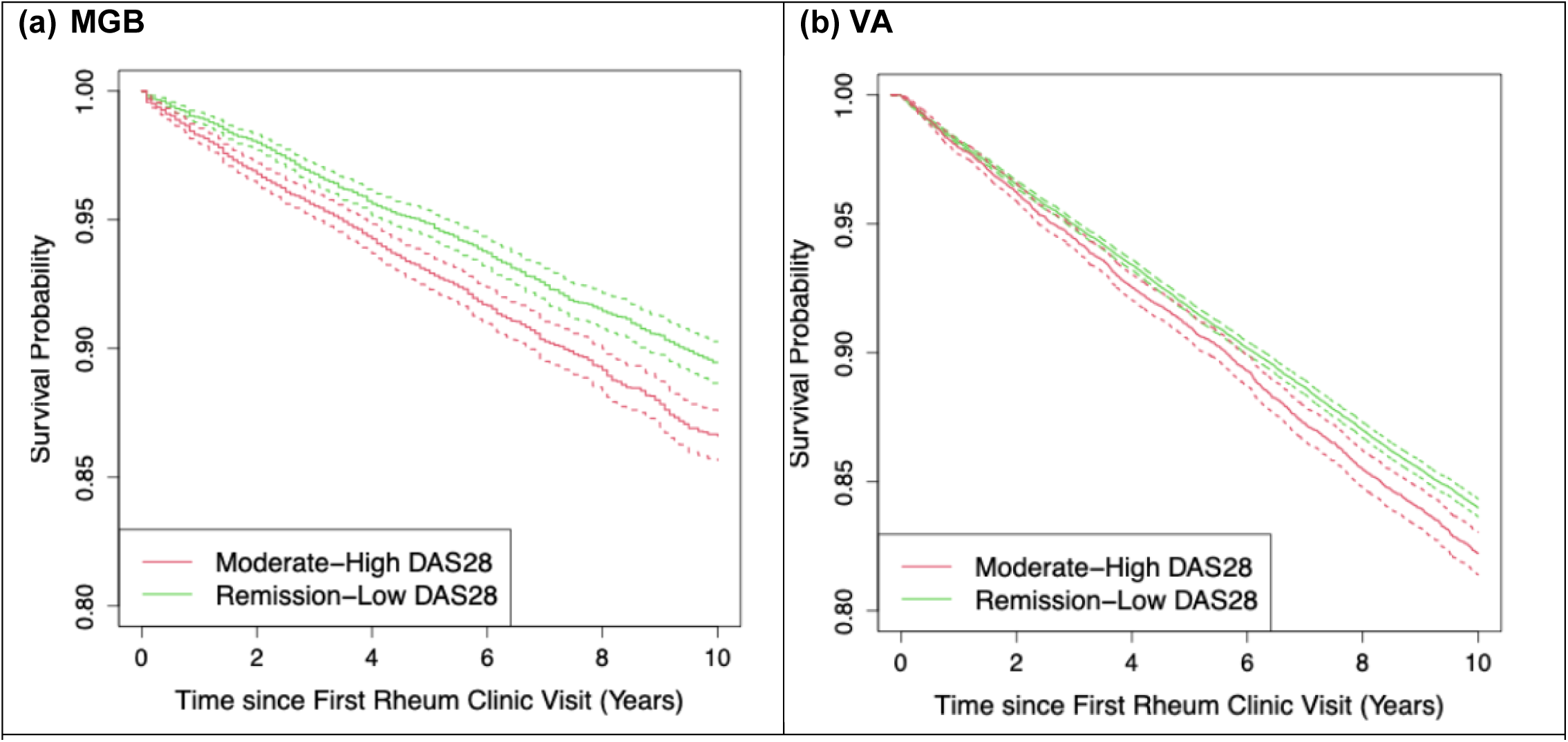
Survival curves with 95% CIs (dotted lines) demonstrating association between moderate/high disease activity vs remission low and incident MACE at (a) MGB and (b) VA, with HR=1.12 (95% CI 1.00-1.25) for MGB and HR=1.14 (1.08-1.21) in VA of inferred moderate-high disease activity for subsequent MACE in Cox analyses, adjusted for baseline covariates and stratified by calendar year.

## Discussion

In this study, we demonstrate that disease activity can be inferred at scale with reasonable performance using data readily available in most EHRs. Comprehensively incorporating longitudinal structured and narrative features from EHRs enabled time-specific inferences about disease activity, which is an essential variable to generate RWE from RWD studies in RA. The framework outlined in this study provides a means to approximate disease activity data over available follow-up in EHR-based cohorts, which would otherwise only be available in prospective longitudinal studies. While the data are sufficiently informative to infer disease activity within an institution, the informativeness of features may vary across institutions leading to lack of transportability of the algorithms across institutions. However, inferred disease activity from models trained within the same institution achieved sufficient accuracy to replicate known associations with subsequent risk of MACE.

As observed in prior studies, models trained using only codified features achieved only modest AUCs with or without the use of machine learning [33–34]. Additionally, incorporating laboratory values into these models led to substantial improvements in their AUCs, despite high rates of missingness at both institutions. We applied simple single-imputation procedures to address missing values, which improved model performance. Inclusion of NLP narrative features to models with codified features led to the largest improvements in the AUC. This finding supports the clinical intuition that the description of a patient’s disease status in the notes provides orthogonal information needed to infer disease activity that are not available in lab results or diagnoses codes [34]. In addition to good discrimination performance reflected in the AUC, models with the full set of structured and narrative data were also able to achieve close agreement between the predicted and observed probabilities of moderate-high disease activity.

Algorithms trained at one institution were not readily transportable to a different institution. The lack of transportability was not surprising given significant differences in patient composition, with older male patients at the VA and predominantly female patients at MGB. Potential differences in the importance of specific codes may be due in part to inherent differences in the healthcare systems. MGB is reimbursed by patients’ private insurance, while Veterans’ healthcare is covered by the VA. Caution is thus warranted in general when transporting models trained at one institution to external institutions.

Ideally, models trained at one institution would be re-trained or calibrated for use at other institutions. The performance of the within-institution algorithms suggests that the model development process itself can generally be repeated at other institutions with a registry linked to EHR to obtain an algorithm with reasonable accuracy, although the vast majority of EHRs won’t have a linked registry.

Using the inferred measures of disease activity, we replicated known associations between higher disease activity and future risk for MACE in large EHR cohorts [35]. This demonstrates how these measures can be used for downstream analyses in large EHR populations in which disease activity at specific time points would otherwise be unavailable. In addition to being an exposure, as in this analysis, these measures can also be used as outcomes or used to define patient populations of interest. Calibration may be warranted to ensure that such analyses are free of bias from use of inferred measures [36].

There were limitations in our study. When incorporating NLP features into our models, we used simple “one-hot” encoding to indicate the presence/absence of terms in proximal narrative notes. Improvements in performance can likely be achieved using word embeddings that represent terms in context [37]. The rapid advancement in large language models (LLMs) also provide opportunities to infer disease activity from EHR features, likely at a much higher cost and need for resources. For example, development of in-house LLMs generally require large datasets, on-premise servers with graphic processing units, Health Insurance Portability and Accountability Act- (HIPAA-) compliant infrastructure, and teams of specialists to develop and maintain the models [38]. Future work will explore the use of LLMs as an option. Our models were trained using data from patients who participated in registry studies, who may differ from the overall RA population at each institution.

These differences could bias the trained models towards disease activity levels observed among sub-populations more likely to participate in these registries. Further validation among broader RA populations at each institution is needed to confirm the performance in the overall populations.

We used DAS28-CRP to measure disease activity in this investigation, as it is a validated measure widely used in clinical studies. Nevertheless, DAS28-CRP may not be fully reflective of disease activity that is observed clinically. Different results may be expected if other measures or criteria are used. While linkage to external claims or death records may partially address this issue, the possibility of missed MACE events outside the VA/MGB systems remains a limitation.

We provide a framework, method, and algorithm to infer disease activity in a manner that can be readily adopted in EHR-based clinical studies, expanding opportunities to study the effect of treatments with large RWD. The methods allow for inference of disease activity at specified time-points throughout a patient’s follow-up. This advancement addresses an important need for studies that require larger populations to study outcomes such as MACE, or studies comparing other safety and effectiveness outcomes, including less common adverse events. Future work includes utilizing inferred disease activity for large-scale trial emulation studies in RA using RWD.

## Data Availability

All data produced in the present work are contained in manuscript and supplementary. Identifiable patient-level data cannot be shared to protect subject privacy.

## Acknowledgements

The opinions expressed in this article are those of the authors and do not necessarily represent those of the Department of Veterans Affairs or the United States government.

