## Supplemental Table 1 for "Inferring rheumatoid arthritis disease activity status from the electronic health records across health systems to enable real-world data studies"

**Supplemental Table 1: Structured data including diagnosis codes, procedure codes, medication codes, and laboratory codes associated with RA and disease activity considered in the disease activity algorithm.**

| **Code** | **Description** | **Code Type** | **Institution** |
| --- | --- | --- | --- |
| CCS:150 | division of joint capsule, ligament or cartilage | CCS | MGB/VA |
| CCS:155 | arthrocentesis | CCS | MGB/VA |
| CCS:183 | routine chest x-ray | CCS | MGB/VA |
| CCS:226 | other diagnostic radiology and related techniques | CCS | MGB/VA |
| LOINC:11090-8 | anti-sm ab (group:sm) | LOINC | MGB |
| LOINC:15205-8 | rheumatoid factor (group:rhf) | LOINC | MGB |
| LOINC:26458-0 | loinc:erythrocytes | LOINC | VA |
| LOINC:29953-7 | ana (quant) (group:anaqn) | LOINC | MGB |
| LOINC:30522-7 | crp, high sens. (cardio) (group:hscrp) | LOINC | MGB |
| LOINC:32218-0 | ccp ab(s) (group:accp) | LOINC | MGB/VA |
| LOINC:33935-8 | ccp ab, igg (group:antccp) | LOINC | MGB |
| LOINC:35492-8 | loinc:staphylococcus aureus.methicillin resistant dna | LOINC | MGB |
| LOINC:4537-7 | esr (group:esr) | LOINC | MGB/VA |
| LOINC:5130-0 | anti-dsdna ab (group:dna) | LOINC | MGB |
| LOINC:5902-2 | loinc:coagulation tissue factor induced | LOINC | MGB/VA |
| LOINC:8061-4 | loinc:nuclear ab | LOINC | MGB |
| LOINC:X1166-8 | crp (mg/l) (group:crpt) | LOINC | MGB |
| PheCode:0086 | viral enteritis | PheCode | MGB/VA |
| PheCode:010 | tuberculosis | PheCode | MGB/VA |
| PheCode:0412 | streptococcus infection | PheCode | MGB/VA |
| PheCode:04121 | rheumatic fever / chorea | PheCode | MGB/VA |
| PheCode:070 | viral hepatitis | PheCode | MGB/VA |
| PheCode:0701 | viral hepatitis a | PheCode | MGB/VA |
| PheCode:0709 | hepatitis nos | PheCode | MGB/VA |
| PheCode:1171 | histoplasmosis | PheCode | VA |
| PheCode:1172 | coccidioidomycosis | PheCode | VA |
| PheCode:200 | myeloproliferative disease | PheCode | MGB/VA |
| PheCode:2044 | multiple myeloma | PheCode | MGB/VA |
| PheCode:244 | hypothyroidism | PheCode | MGB/VA |
| PheCode:245 | thyroiditis | PheCode | MGB/VA |
| PheCode:25521 | glucocorticoid deficiency | PheCode | MGB/VA |
| PheCode:261 | vitamin deficiency | PheCode | MGB/VA |
| PheCode:2614 | vitamin d deficiency | PheCode | MGB/VA |
| PheCode:26141 | rickets or osteomalacia | PheCode | VA |
| PheCode:27031 | polyclonal hypergammaglobulinemia | PheCode | MGB/VA |
| PheCode:27033 | amyloidosis | PheCode | MGB/VA |
| PheCode:274 | gout and other crystal arthropathies | PheCode | MGB/VA |
| PheCode:2741 | gout | PheCode | MGB/VA |
| PheCode:27411 | gouty arthropathy | PheCode | MGB/VA |
| PheCode:2742 | crystal arthropathies | PheCode | MGB/VA |
| PheCode:27421 | chondrocalcinosis | PheCode | MGB/VA |
| PheCode:279 | disorders involving the immune mechanism | PheCode | MGB/VA |
| PheCode:2791 | immunity deficiency | PheCode | MGB/VA |
| PheCode:2792 | autoimmune disease nec | PheCode | MGB/VA |
| PheCode:28611 | von willebrand's disease | PheCode | MGB/VA |
| PheCode:2872 | allergic purpura | PheCode | VA |
| PheCode:2891 | myelofibrosis | PheCode | VA |
| PheCode:2894 | lymphadenitis | PheCode | MGB/VA |
| PheCode:29016 | vascular dementia | PheCode | MGB/VA |
| PheCode:30331 | gastrointestinal malfunction arising from mental factors | PheCode | MGB/VA |
| PheCode:335 | multiple sclerosis | PheCode | MGB/VA |
| PheCode:3592 | myopathy | PheCode | MGB/VA |
| PheCode:3625 | toxic maculopathy of retina | PheCode | MGB/VA |
| PheCode:370 | keratitis | PheCode | MGB/VA |
| PheCode:3703 | keratoconjunctivitis | PheCode | MGB/VA |
| PheCode:37031 | keratoconjunctivitis sicca | PheCode | MGB/VA |
| PheCode:371 | inflammation of the eye | PheCode | MGB/VA |
| PheCode:3711 | uveitis, noninfectious or nos | PheCode | MGB/VA |
| PheCode:37121 | allergic conjunctivitis | PheCode | MGB/VA |
| PheCode:3713 | inflammation of eyelids | PheCode | MGB/VA |
| PheCode:3719 | chronic inflammatory disorders of orbit | PheCode | VA |
| PheCode:3751 | dry eyes | PheCode | MGB/VA |
| PheCode:3791 | scleritis and episcleritis | PheCode | MGB/VA |
| PheCode:3811 | otitis media | PheCode | MGB/VA |
| PheCode:3813 | mastoiditis & related conditions | PheCode | MGB/VA |
| PheCode:3841 | myringitis | PheCode | MGB/VA |
| PheCode:3863 | labyrinthitis | PheCode | MGB/VA |
| PheCode:411 | ischemic heart disease | PheCode | MGB/VA |
| PheCode:420 | carditis | PheCode | MGB/VA |
| PheCode:4201 | myocarditis | PheCode | MGB/VA |
| PheCode:4202 | pericarditis | PheCode | MGB/VA |
| PheCode:4203 | endocarditis | PheCode | MGB/VA |
| PheCode:4431 | raynaud's syndrome | PheCode | MGB/VA |
| PheCode:446 | polyarteritis nodosa and allied conditions | PheCode | MGB/VA |
| PheCode:4463 | hypersensitivity angiitis | PheCode | MGB/VA |
| PheCode:4464 | wegener's granulomatosis | PheCode | MGB/VA |
| PheCode:4465 | giant cell arteritis | PheCode | MGB/VA |
| PheCode:4466 | polyarteritis nodosa | PheCode | MGB/VA |
| PheCode:4469 | arteritis nos | PheCode | MGB/VA |
| PheCode:476 | allergic rhinitis | PheCode | MGB/VA |
| PheCode:481 | influenza | PheCode | MGB/VA |
| PheCode:495 | asthma | PheCode | MGB/VA |
| PheCode:497 | bronchitis | PheCode | MGB/VA |
| PheCode:499 | cystic fibrosis | PheCode | MGB/VA |
| PheCode:5002 | pneumoconiosis | PheCode | MGB/VA |
| PheCode:502 | postinflammatory pulmonary fibrosis | PheCode | MGB/VA |
| PheCode:5041 | idiopathic fibrosing alveolitis | PheCode | MGB/VA |
| PheCode:510 | other diseases of lung | PheCode | MGB/VA |
| PheCode:5127 | shortness of breath | PheCode | MGB/VA |
| PheCode:5128 | cough | PheCode | MGB/VA |
| PheCode:521 | diseases of hard tissues of teeth | PheCode | MGB/VA |
| PheCode:5231 | gingivitis | PheCode | MGB/VA |
| PheCode:5264 | temporomandibular joint disorders | PheCode | MGB/VA |
| PheCode:5265 | inflammatory conditions of jaw | PheCode | MGB/VA |
| PheCode:5271 | hypertrophy of salivary gland | PheCode | VA |
| PheCode:5272 | sialoadenitis | PheCode | MGB/VA |
| PheCode:5291 | glossitis | PheCode | MGB/VA |
| PheCode:5301 | esophagitis, gerd and related diseases | PheCode | MGB/VA |
| PheCode:53011 | gerd | PheCode | MGB/VA |
| PheCode:53014 | reflux esophagitis | PheCode | MGB/VA |
| PheCode:5309 | heartburn | PheCode | MGB/VA |
| PheCode:5315 | gastrojejunal ulcer | PheCode | VA |
| PheCode:5356 | duodenitis | PheCode | MGB/VA |
| PheCode:5401 | appendicitis | PheCode | MGB/VA |
| PheCode:555 | inflammatory bowel disease and other gastroenteritis and colitis | PheCode | MGB/VA |
| PheCode:5551 | regional enteritis | PheCode | MGB/VA |
| PheCode:5552 | ulcerative colitis | PheCode | MGB/VA |
| PheCode:5611 | diarrhea | PheCode | MGB/VA |
| PheCode:5621 | diverticulosis | PheCode | MGB/VA |
| PheCode:5622 | diverticulitis | PheCode | MGB/VA |
| PheCode:5641 | irritable bowel syndrome | PheCode | MGB/VA |
| PheCode:5801 | glomerulonephritis | PheCode | MGB/VA |
| PheCode:5804 | renal sclerosis, nos | PheCode | MGB/VA |
| PheCode:590 | pyelonephritis | PheCode | MGB/VA |
| PheCode:5921 | cystitis | PheCode | MGB/VA |
| PheCode:5922 | urethritis and urethral syndrome | PheCode | MGB/VA |
| PheCode:5993 | dysuria | PheCode | MGB/VA |
| PheCode:601 | inflammatory diseases of prostate | PheCode | MGB/VA |
| PheCode:6011 | prostatitis | PheCode | MGB/VA |
| PheCode:6131 | inflammatory disease of breast | PheCode | MGB/VA |
| PheCode:6143 | pelvic inflammatory disease (pid) | PheCode | MGB/VA |
| PheCode:61433 | pelvic inflammatory disease, nos | PheCode | MGB/VA |
| PheCode:6865 | pyoderma | PheCode | VA |
| PheCode:69521 | dermatitis herpetiformis | PheCode | VA |
| PheCode:6954 | lupus (localized and systemic) | PheCode | MGB/VA |
| PheCode:69542 | systemic lupus erythematosus | PheCode | MGB/VA |
| PheCode:69581 | erythema nodosum | PheCode | MGB |
| PheCode:696 | psoriasis and related disorders | PheCode | MGB/VA |
| PheCode:6964 | psoriasis | PheCode | MGB/VA |
| PheCode:69641 | psoriasis vulgaris | PheCode | MGB/VA |
| PheCode:69642 | psoriatic arthropathy | PheCode | MGB/VA |
| PheCode:697 | sarcoidosis | PheCode | MGB/VA |
| PheCode:7053 | hidradenitis | PheCode | MGB/VA |
| PheCode:709 | diffuse diseases of connective tissue | PheCode | MGB/VA |
| PheCode:7092 | sicca syndrome | PheCode | MGB/VA |
| PheCode:7093 | systemic sclerosis | PheCode | MGB/VA |
| PheCode:7094 | polymyositis | PheCode | MGB/VA |
| PheCode:7095 | dermatomyositis | PheCode | MGB/VA |
| PheCode:7096 | other specified diffuse diseases of connective tissue | PheCode | MGB/VA |
| PheCode:7097 | unspecified diffuse connective tissue disease | PheCode | MGB/VA |
| PheCode:71019 | unspecified osteomyelitis | PheCode | MGB/VA |
| PheCode:711 | arthropathy associated with infections | PheCode | MGB/VA |
| PheCode:7111 | pyogenic arthritis | PheCode | MGB/VA |
| PheCode:7112 | reiter's disease | PheCode | MGB/VA |
| PheCode:713 | arthropathy associated with other disorders classified elsewhere | PheCode | MGB/VA |
| PheCode:7135 | arthropathy associated with neurological disorders | PheCode | VA |
| PheCode:714 | rheumatoid arthritis and other inflammatory polyarthropathies | PheCode | MGB/VA |
| PheCode:7141 | rheumatoid arthritis | PheCode | MGB/VA |
| PheCode:7142 | juvenile rheumatoid arthritis | PheCode | MGB/VA |
| PheCode:715 | other inflammatory spondylopathies | PheCode | MGB/VA |
| PheCode:7151 | sacroiliitis nec | PheCode | MGB/VA |
| PheCode:7152 | ankylosing spondylitis | PheCode | MGB/VA |
| PheCode:716 | other arthropathies | PheCode | MGB/VA |
| PheCode:7161 | unspecified polyarthropathy or polyarthritis | PheCode | MGB/VA |
| PheCode:7162 | unspecified monoarthritis | PheCode | MGB/VA |
| PheCode:7163 | kaschin-beck disease | PheCode | VA |
| PheCode:7168 | palindromic rheumatism | PheCode | MGB/VA |
| PheCode:7169 | arthropathy nos | PheCode | MGB/VA |
| PheCode:717 | polymyalgia rheumatica | PheCode | MGB/VA |
| PheCode:726 | peripheral enthesopathies and allied syndromes | PheCode | MGB/VA |
| PheCode:7261 | enthesopathy | PheCode | MGB/VA |
| PheCode:7262 | synoviopathy | PheCode | MGB/VA |
| PheCode:7263 | bursitis | PheCode | MGB/VA |
| PheCode:727 | other disorders of synovium, tendon, and bursa | PheCode | MGB/VA |
| PheCode:7271 | synovitis and tenosynovitis | PheCode | MGB/VA |
| PheCode:7272 | bursitis disorders | PheCode | MGB/VA |
| PheCode:7275 | rupture of synovium | PheCode | MGB/VA |
| PheCode:7277 | contracture of tendon (sheath) | PheCode | MGB/VA |
| PheCode:728 | disorders of muscle, ligament, and fascia | PheCode | MGB/VA |
| PheCode:7287 | fasciitis | PheCode | MGB/VA |
| PheCode:7291 | rheumatism, unspecified and fibrositis | PheCode | MGB/VA |
| PheCode:7293 | panniculitis | PheCode | MGB/VA |
| PheCode:7311 | osteitis deformans [paget's disease of bone] | PheCode | MGB/VA |
| PheCode:7336 | costochondritis | PheCode | MGB/VA |
| PheCode:73521 | hammer toe (acquired) | PheCode | MGB/VA |
| PheCode:73523 | hallux rigidus | PheCode | MGB/VA |
| PheCode:7353 | hallux valgus (bunion) | PheCode | MGB/VA |
| PheCode:7371 | kyphosis (acquired) | PheCode | MGB/VA |
| PheCode:739 | contracture of joint | PheCode | MGB/VA |
| PheCode:740 | osteoarthrosis | PheCode | MGB/VA |
| PheCode:7401 | osteoarthritis; localized | PheCode | MGB/VA |
| PheCode:74011 | osteoarthrosis, localized, primary | PheCode | MGB/VA |
| PheCode:74012 | osteoarthrosis, localized, secondary | PheCode | MGB/VA |
| PheCode:7402 | osteoarthrosis, generalized | PheCode | MGB/VA |
| PheCode:7403 | osteoarthrosis involving more than one site, but not specified as generalized | PheCode | MGB/VA |
| PheCode:7409 | osteoarthrosis nos | PheCode | MGB/VA |
| PheCode:741 | symptoms and disorders of the joints | PheCode | MGB/VA |
| PheCode:7411 | ankylosis of joint | PheCode | MGB/VA |
| PheCode:7412 | stiffness of joint | PheCode | MGB/VA |
| PheCode:7414 | joint effusions | PheCode | MGB/VA |
| PheCode:7415 | hemarthrosis | PheCode | MGB/VA |
| PheCode:7416 | villonodular synovitis | PheCode | MGB/VA |
| PheCode:7421 | loose body in joint | PheCode | MGB/VA |
| PheCode:7428 | articular cartilage disorder | PheCode | MGB/VA |
| PheCode:7431 | osteoporosis | PheCode | MGB/VA |
| PheCode:74311 | osteoporosis nos | PheCode | MGB/VA |
| PheCode:74312 | senile osteoporosis | PheCode | MGB/VA |
| PheCode:74313 | other specified osteoporosis | PheCode | MGB/VA |
| PheCode:7434 | stress fracture | PheCode | MGB/VA |
| PheCode:7439 | osteopenia or other disorder of bone and cartilage | PheCode | MGB/VA |
| PheCode:745 | pain in joint | PheCode | MGB/VA |
| PheCode:770 | myalgia and myositis unspecified | PheCode | MGB/VA |
| PheCode:773 | pain in limb | PheCode | MGB/VA |
| PheCode:7823 | edema | PheCode | MGB/VA |
| PheCode:783 | fever of unknown origin | PheCode | MGB/VA |
| PheCode:7901 | elevated sedimentation rate | PheCode | MGB/VA |
| PheCode:7908 | elevated c-reactive protein (crp) | PheCode | MGB/VA |
| PheCode:791 | gangrene | PheCode | MGB/VA |
| PheCode:798 | malaise and fatigue | PheCode | MGB/VA |
| PheCode:7981 | chronic fatigue syndrome | PheCode | MGB/VA |
| PheCode:836 | traumatic arthropathy | PheCode | MGB/VA |
| PheCode:947 | urticaria | PheCode | MGB/VA |
| RXNORM:1227 | auranofin | RXNORM | MGB/VA |
| RXNORM:1357536 | tofacitinib | RXNORM | MGB/VA |
| RXNORM:1599788 | secukinumab | RXNORM | VA |
| RXNORM:191831 | infliximab | RXNORM | MGB/VA |
| RXNORM:214555 | etanercept | RXNORM | MGB/VA |
| RXNORM:27169 | leflunomide | RXNORM | MGB/VA |
| RXNORM:32613 | oxaprozin | RXNORM | MGB/VA |
| RXNORM:327361 | adalimumab | RXNORM | MGB/VA |
| RXNORM:3355 | diclofenac | RXNORM | MGB/VA |
| RXNORM:3393 | diflunisal | RXNORM | MGB/VA |
| RXNORM:41493 | meloxicam | RXNORM | MGB/VA |
| RXNORM:46041 | alendronate | RXNORM | MGB/VA |
| RXNORM:5224 | heparin | RXNORM | MGB/VA |
| RXNORM:5521 | hydroxychloroquine | RXNORM | MGB/VA |
| RXNORM:6038 | isoniazid | RXNORM | MGB/VA |
| RXNORM:612865 | tocilizumab | RXNORM | MGB/VA |
| RXNORM:614391 | abatacept | RXNORM | MGB/VA |
| RXNORM:6851 | methotrexate | RXNORM | MGB/VA |
| RXNORM:709271 | certolizumab | RXNORM | MGB/VA |
| RXNORM:72435 | anakinra | RXNORM | MGB/VA |
| RXNORM:7258 | naproxen | RXNORM | MGB/VA |
| RXNORM:73056 | risedronate | RXNORM | MGB/VA |
| RXNORM:787390 | tapentadol | RXNORM | MGB/VA |
| RXNORM:797195 | fesoterodine | RXNORM | MGB/VA |
| RXNORM:819300 | golimumab | RXNORM | MGB/VA |
| RXNORM:8640 | prednisone | RXNORM | MGB/VA |
| RXNORM:9524 | sulfasalazine | RXNORM | MGB/VA |
| ShortName:BUN - BSP | bun - bsp | ShortName | VA |
| ShortName:Baso - Fra | baso - fra | ShortName | VA |
| ShortName:CCP | ccp | ShortName | VA |
| ShortName:CRP | crp | ShortName | VA |
| ShortName:Creat - BSP | creat - bsp | ShortName | VA |
| ShortName:ESR | esr | ShortName | VA |
| ShortName:Eos - Abs | eos - abs | ShortName | VA |
| ShortName:Eos - Fra | eos - fra | ShortName | VA |
| ShortName:HCT | hct | ShortName | VA |
| ShortName:HLAB27 | hlab27 | ShortName | VA |
| ShortName:Hemoglobin | hemoglobin | ShortName | VA |
| ShortName:INR | inr | ShortName | VA |
| ShortName:Lymph - Fra | lymph - fra | ShortName | VA |
| ShortName:MCH | mch | ShortName | VA |
| ShortName:MCHC | mchc | ShortName | VA |
| ShortName:MCV | mcv | ShortName | VA |
| ShortName:MPV | mpv | ShortName | VA |
| ShortName:Mg - BSP | mg - bsp | ShortName | VA |
| ShortName:Mono - Fra | mono - fra | ShortName | VA |
| ShortName:Neut - Abs | neut - abs | ShortName | VA |
| ShortName:Neut - Fra | neut - fra | ShortName | VA |
| ShortName:Platelet | platelet | ShortName | VA |
| ShortName:Potas - BSP | potas - bsp | ShortName | VA |
| ShortName:RBC | rbc | ShortName | VA |
| ShortName:RDW | rdw | ShortName | VA |
| ShortName:RF | rf | ShortName | VA |
| ShortName:WBC | wbc | ShortName | VA |
