## Supplemental Table 2 for "Inferring rheumatoid arthritis disease activity status from the electronic health records across health systems to enable real-world data studies"

**Supplemental Table 2: Narrative features represented as concept unique identifiers (CUIs) related to RA and disease activity considered in the disease activity algorithm.**

| **Code** | **Description** | **Code Type** | **Institution** | **Code Source** |
| --- | --- | --- | --- | --- |
| C0000768 | Congenital Abnormality | CUI | MGB/VA | ONCE |
| C0000970 | acetaminophen | CUI | MGB/VA | ONCE |
| C0001347 | Acute-Phase Proteins | CUI | MGB/VA | ONCE |
| C0001807 | Aggressive behavior | CUI | MGB/VA | DAS Manual Curation |
| C0002600 | amitriptyline | CUI | MGB/VA | DAS Manual Curation |
| C0002716 | Amyloid | CUI | MGB/VA | DAS Manual Curation |
| C0002726 | Amyloidosis | CUI | MGB/VA | ONCE |
| C0002766 | Pain management | CUI | MGB/VA | ONCE |
| C0002771 | Analgesics | CUI | MGB/VA | ONCE |
| C0002871 | Anemia | CUI | MGB/VA | ONCE |
| C0002873 | Anemia of chronic disease | CUI | MGB/VA | ONCE |
| C0003191 | Antirheumatic Agents | CUI | MGB/VA | DAS Manual Curation |
| C0003209 | Anti-Inflammatory Agents | CUI | MGB/VA | DAS Manual Curation |
| C0003211 | Anti-Inflammatory Agents, Non-Steroidal | CUI | MGB/VA | ONCE |
| C0003232 | Antibiotics | CUI | MGB/VA | ONCE |
| C0003237 | Antibiotics, Antitubercular | CUI | MGB/VA | DAS Manual Curation |
| C0003241 | Antibodies | CUI | MGB/VA | DAS Manual Curation |
| C0003243 | Antibodies, Antinuclear | CUI | MGB/VA | ONCE |
| C0003850 | Arteriosclerosis | CUI | MGB/VA | ONCE |
| C0003862 | Arthralgia | CUI | MGB/VA | ONCE |
| C0003864 | Arthritis | CUI | MGB/VA | ONCE |
| C0003868 | Arthritis, Gouty | CUI | MGB/VA | ONCE |
| C0003869 | Arthritis, Infectious | CUI | MGB/VA | ONCE |
| C0003872 | Arthritis, Psoriatic | CUI | MGB/VA | ONCE |
| C0003873 | Rheumatoid Arthritis | CUI | MGB/VA | ONCE |
| C0003893 | Arthroplasty | CUI | MGB/VA | ONCE |
| C0004093 | Asthenia | CUI | MGB/VA | ONCE |
| C0004096 | Asthma | CUI | MGB/VA | ONCE |
| C0004364 | Autoimmune Diseases | CUI | MGB/VA | ONCE |
| C0004368 | Autoimmune state | CUI | MGB/VA | ONCE |
| C0004482 | azathioprine | CUI | MGB/VA | DAS Manual Curation |
| C0004604 | Back Pain | CUI | MGB/VA | DAS Manual Curation |
| C0004951 | Beliefs | CUI | MGB/VA | DAS Manual Curation |
| C0005437 | Bilirubin | CUI | MGB/VA | ONCE |
| C0005515 | Biological Factors | CUI | MGB/VA | DAS Manual Curation |
| C0005527 | Biological Response Modifier Therapy | CUI | MGB/VA | ONCE |
| C0005558 | Biopsy | CUI | MGB/VA | ONCE |
| C0005824 | Blood pressure determination | CUI | MGB/VA | DAS Manual Curation |
| C0005889 | Body Fluids | CUI | MGB/VA | DAS Manual Curation |
| C0006444 | Bursitis | CUI | MGB/VA | DAS Manual Curation |
| C0006560 | C-reactive protein | CUI | MGB/VA | ONCE |
| C0006675 | calcium | CUI | MGB/VA | DAS Manual Curation |
| C0006726 | Calcium, Dietary | CUI | MGB/VA | ONCE |
| C0007584 | Cell Count | CUI | MGB/VA | DAS Manual Curation |
| C0007642 | Cellulitis | CUI | MGB/VA | ONCE |
| C0008269 | chloroquine | CUI | MGB/VA | DAS Manual Curation |
| C0008300 | Choice Behavior | CUI | MGB/VA | DAS Manual Curation |
| C0008679 | Chronic disease | CUI | MGB/VA | ONCE |
| C0009324 | Ulcerative Colitis | CUI | MGB/VA | ONCE |
| C0009326 | Collagen Diseases | CUI | MGB/VA | ONCE |
| C0009429 | Combined Modality Therapy | CUI | MGB/VA | DAS Manual Curation |
| C0009450 | Communicable Diseases | CUI | MGB/VA | ONCE |
| C0009566 | Complication | CUI | MGB/VA | ONCE |
| C0009676 | Confusion | CUI | MGB/VA | ONCE |
| C0009763 | Conjunctivitis | CUI | MGB/VA | ONCE |
| C0009782 | Connective Tissue Diseases | CUI | MGB/VA | ONCE |
| C0010068 | Coronary heart disease | CUI | MGB/VA | ONCE |
| C0010294 | creatinine | CUI | MGB/VA | DAS Manual Curation |
| C0010583 | cyclophosphamide | CUI | MGB/VA | DAS Manual Curation |
| C0010592 | cyclosporine | CUI | MGB/VA | DAS Manual Curation |
| C0010957 | Tissue damage | CUI | MGB/VA | ONCE |
| C0011164 | Abnormal degeneration | CUI | MGB/VA | ONCE |
| C0011560 | Amyloid deposition | CUI | MGB/VA | DAS Manual Curation |
| C0011603 | Dermatitis | CUI | MGB/VA | ONCE |
| C0011644 | Scleroderma | CUI | MGB/VA | ONCE |
| C0011777 | dexamethasone | CUI | MGB/VA | DAS Manual Curation |
| C0011847 | Diabetes | CUI | MGB/VA | ONCE |
| C0011923 | Diagnostic Imaging | CUI | MGB/VA | ONCE |
| C0011991 | Diarrhea | CUI | MGB/VA | ONCE |
| C0012619 | disc disorder | CUI | MGB/VA | DAS Manual Curation |
| C0012624 | Discitis | CUI | MGB/VA | ONCE |
| C0012634 | Disease | CUI | MGB/VA | ONCE |
| C0012691 | Dislocations | CUI | MGB/VA | DAS Manual Curation |
| C0013216 | Pharmacotherapy | CUI | MGB/VA | DAS Manual Curation |
| C0013220 | Drug Tolerance | CUI | MGB/VA | DAS Manual Curation |
| C0013221 | Drug toxicity | CUI | MGB/VA | DAS Manual Curation |
| C0013227 | Pharmaceutical Preparations | CUI | MGB/VA | DAS Manual Curation |
| C0013238 | Dry Eye Syndromes | CUI | MGB/VA | ONCE |
| C0013274 | Patent ductus arteriosus | CUI | MGB/VA | ONCE |
| C0013604 | Edema | CUI | MGB/VA | ONCE |
| C0013687 | effusion | CUI | MGB/VA | ONCE |
| C0015230 | Exanthema | CUI | MGB/VA | ONCE |
| C0015672 | Fatigue | CUI | MGB/VA | ONCE |
| C0015674 | Chronic Fatigue Syndrome | CUI | MGB/VA | ONCE |
| C0015967 | Fever | CUI | MGB/VA | ONCE |
| C0016053 | Fibromyalgia | CUI | MGB/VA | ONCE |
| C0016059 | Fibrosis | CUI | MGB/VA | ONCE |
| C0016286 | Fluid Therapy | CUI | MGB/VA | ONCE |
| C0016410 | folic acid | CUI | MGB/VA | ONCE |
| C0016506 | Foot Deformities | CUI | MGB/VA | DAS Manual Curation |
| C0016512 | Foot pain | CUI | MGB/VA | DAS Manual Curation |
| C0017925 | Glycogen Storage Disease Type VI | CUI | MGB/VA | DAS Manual Curation |
| C0017968 | Glycoproteins | CUI | MGB/VA | ONCE |
| C0018033 | aurothioglucose | CUI | MGB/VA | DAS Manual Curation |
| C0018592 | Happiness | CUI | MGB/VA | DAS Manual Curation |
| C0018801 | Heart failure | CUI | MGB/VA | ONCE |
| C0018802 | Congestive heart failure | CUI | MGB/VA | ONCE |
| C0018862 | Heberden node | CUI | MGB/VA | ONCE |
| C0018941 | Hematologic Tests | CUI | MGB/VA | ONCE |
| C0018995 | Hemochromatosis | CUI | MGB/VA | ONCE |
| C0019158 | Hepatitis | CUI | MGB/VA | ONCE |
| C0019159 | Hepatitis A | CUI | MGB/VA | ONCE |
| C0019163 | Hepatitis B | CUI | MGB/VA | ONCE |
| C0019196 | Hepatitis C | CUI | MGB/VA | ONCE |
| C0019559 | Hip joint pain | CUI | MGB/VA | DAS Manual Curation |
| C0020311 | Hydrotherapy | CUI | MGB/VA | DAS Manual Curation |
| C0020336 | hydroxychloroquine | CUI | MGB/VA | ONCE |
| C0020492 | Hyperostosis | CUI | MGB/VA | DAS Manual Curation |
| C0020517 | Hypersensitivity | CUI | MGB/VA | ONCE |
| C0020538 | Hypertensive disease | CUI | MGB/VA | ONCE |
| C0020542 | Pulmonary Hypertension | CUI | MGB/VA | ONCE |
| C0020564 | Hypertrophy | CUI | MGB/VA | ONCE |
| C0020740 | ibuprofen | CUI | MGB/VA | DAS Manual Curation |
| C0020963 | Immune Tolerance | CUI | MGB/VA | DAS Manual Curation |
| C0020971 | Immunization | CUI | MGB/VA | ONCE |
| C0021053 | Immune System Diseases | CUI | MGB/VA | ONCE |
| C0021079 | Therapeutic immunosuppression | CUI | MGB/VA | DAS Manual Curation |
| C0021081 | Immunosuppressive Agents | CUI | MGB/VA | DAS Manual Curation |
| C0021368 | Inflammation | CUI | MGB/VA | ONCE |
| C0021376 | Chronic inflammation | CUI | MGB/VA | ONCE |
| C0021386 | Necrotizing granulomatous inflammation | CUI | MGB/VA | ONCE |
| C0021390 | Inflammatory Bowel Diseases | CUI | MGB/VA | ONCE |
| C0021440 | Intravenous infusion procedures | CUI | MGB/VA | ONCE |
| C0021485 | Injection of therapeutic agent | CUI | MGB/VA | ONCE |
| C0021943 | Chromosome inversion | CUI | MGB/VA | DAS Manual Curation |
| C0021945 | Body part inversion | CUI | MGB/VA | DAS Manual Curation |
| C0022081 | Iritis | CUI | MGB/VA | ONCE |
| C0022408 | Arthropathy | CUI | MGB/VA | ONCE |
| C0022410 | Joint Instability | CUI | MGB/VA | ONCE |
| C0022658 | Kidney Diseases | CUI | MGB/VA | ONCE |
| C0022885 | Laboratory Procedures | CUI | MGB/VA | DAS Manual Curation |
| C0023343 | Leprosy | CUI | MGB/VA | ONCE |
| C0023413 | leucovorin | CUI | MGB/VA | DAS Manual Curation |
| C0023508 | White Blood Cell Count procedure | CUI | MGB/VA | ONCE |
| C0023518 | Leukocytosis | CUI | MGB/VA | ONCE |
| C0023530 | Leukopenia | CUI | MGB/VA | DAS Manual Curation |
| C0023660 | lidocaine | CUI | MGB/VA | DAS Manual Curation |
| C0024031 | Low Back Pain | CUI | MGB/VA | DAS Manual Curation |
| C0024115 | Lung diseases | CUI | MGB/VA | ONCE |
| C0024131 | Lupus Vulgaris | CUI | MGB/VA | ONCE |
| C0024138 | Lupus Erythematosus, Discoid | CUI | MGB/VA | ONCE |
| C0024141 | Lupus Erythematosus, Systemic | CUI | MGB/VA | ONCE |
| C0024299 | Lymphoma | CUI | MGB/VA | ONCE |
| C0025677 | methotrexate | CUI | MGB/VA | ONCE |
| C0025815 | methylprednisolone | CUI | MGB/VA | DAS Manual Curation |
| C0026272 | Mixed Connective Tissue Disease | CUI | MGB/VA | ONCE |
| C0026837 | Muscle Rigidity | CUI | MGB/VA | DAS Manual Curation |
| C0026846 | Muscular Atrophy | CUI | MGB/VA | ONCE |
| C0026857 | Musculoskeletal Diseases | CUI | MGB/VA | ONCE |
| C0026936 | Mycoplasma Infections | CUI | MGB/VA | ONCE |
| C0027051 | Myocardial Infarction | CUI | MGB/VA | ONCE |
| C0027059 | Myocarditis | CUI | MGB/VA | ONCE |
| C0027396 | naproxen | CUI | MGB/VA | DAS Manual Curation |
| C0027415 | Narcotics | CUI | MGB/VA | DAS Manual Curation |
| C0027497 | Nausea | CUI | MGB/VA | ONCE |
| C0027498 | Nausea and vomiting | CUI | MGB/VA | ONCE |
| C0027627 | Neoplasm Metastasis | CUI | MGB/VA | ONCE |
| C0027707 | Nephritis, Interstitial | CUI | MGB/VA | ONCE |
| C0027947 | Neutropenia | CUI | MGB/VA | ONCE |
| C0028259 | Nodule | CUI | MGB/VA | ONCE |
| C0028678 | nursing therapy | CUI | MGB/VA | ONCE |
| C0028778 | Obstruction | CUI | MGB/VA | ONCE |
| C0029408 | Degenerative polyarthritis | CUI | MGB/VA | ONCE |
| C0029456 | Osteoporosis | CUI | MGB/VA | ONCE |
| C0030193 | Pain | CUI | MGB/VA | ONCE |
| C0030247 | Palpation | CUI | MGB/VA | ONCE |
| C0030554 | Paresthesia | CUI | MGB/VA | DAS Manual Curation |
| C0030695 | Patient Monitoring | CUI | MGB/VA | DAS Manual Curation |
| C0030817 | penicillamine | CUI | MGB/VA | DAS Manual Curation |
| C0030920 | Peptic Ulcer | CUI | MGB/VA | ONCE |
| C0031046 | Pericarditis | CUI | MGB/VA | ONCE |
| C0031350 | Pharyngitis | CUI | MGB/VA | ONCE |
| C0031809 | Physical Examination | CUI | MGB/VA | DAS Manual Curation |
| C0031843 | physiological aspects | CUI | MGB/VA | DAS Manual Curation |
| C0031990 | piroxicam | CUI | MGB/VA | DAS Manual Curation |
| C0032181 | Platelet Count measurement | CUI | MGB/VA | DAS Manual Curation |
| C0032227 | Pleural effusion disorder | CUI | MGB/VA | ONCE |
| C0032231 | Pleurisy | CUI | MGB/VA | ONCE |
| C0032285 | Pneumonia | CUI | MGB/VA | DAS Manual Curation |
| C0032533 | Polymyalgia Rheumatica | CUI | MGB/VA | ONCE |
| C0032952 | prednisone | CUI | MGB/VA | ONCE |
| C0033095 | Pressure- physical agent | CUI | MGB/VA | DAS Manual Curation |
| C0033213 | Problem | CUI | MGB/VA | DAS Manual Curation |
| C0033554 | Prostaglandins | CUI | MGB/VA | ONCE |
| C0033684 | Proteins | CUI | MGB/VA | ONCE |
| C0033774 | Pruritus | CUI | MGB/VA | ONCE |
| C0033802 | Pseudogout | CUI | MGB/VA | ONCE |
| C0033860 | Psoriasis | CUI | MGB/VA | ONCE |
| C0034107 | Pulse taking | CUI | MGB/VA | DAS Manual Curation |
| C0034897 | Recurrence | CUI | MGB/VA | DAS Manual Curation |
| C0034991 | Rehabilitation therapy | CUI | MGB/VA | ONCE |
| C0035139 | Surgical Replantation | CUI | MGB/VA | ONCE |
| C0035204 | Respiration Disorders | CUI | MGB/VA | ONCE |
| C0035435 | Rheumatism | CUI | MGB/VA | ONCE |
| C0035436 | Rheumatic Fever | CUI | MGB/VA | ONCE |
| C0035448 | Rheumatoid Factor | CUI | MGB/VA | ONCE |
| C0035450 | Rheumatoid Nodule | CUI | MGB/VA | ONCE |
| C0035455 | Rhinitis | CUI | MGB/VA | ONCE |
| C0035920 | Rubella | CUI | MGB/VA | ONCE |
| C0036078 | sulfasalazine | CUI | MGB/VA | ONCE |
| C0036202 | Sarcoidosis | CUI | MGB/VA | ONCE |
| C0036396 | Sciatica | CUI | MGB/VA | DAS Manual Curation |
| C0036421 | Systemic Scleroderma | CUI | MGB/VA | ONCE |
| C0037011 | Shoulder Pain | CUI | MGB/VA | DAS Manual Curation |
| C0037369 | Smoking | CUI | MGB/VA | ONCE |
| C0037580 | Soft tissue swelling | CUI | MGB/VA | DAS Manual Curation |
| C0038013 | Ankylosing spondylitis | CUI | MGB/VA | ONCE |
| C0038317 | Steroids | CUI | MGB/VA | ONCE |
| C0038362 | Stomatitis | CUI | MGB/VA | ONCE |
| C0038395 | Streptococcal Infections | CUI | MGB/VA | ONCE |
| C0038435 | Stress | CUI | MGB/VA | DAS Manual Curation |
| C0038999 | Swelling | CUI | MGB/VA | ONCE |
| C0039082 | Syndrome | CUI | MGB/VA | ONCE |
| C0039103 | Synovitis | CUI | MGB/VA | ONCE |
| C0039483 | Giant Cell Arteritis | CUI | MGB/VA | ONCE |
| C0039520 | Tenosynovitis | CUI | MGB/VA | DAS Manual Curation |
| C0039869 | Thinking, function | CUI | MGB/VA | DAS Manual Curation |
| C0039985 | Plain chest X-ray | CUI | MGB/VA | ONCE |
| C0040134 | thyroid (USP) | CUI | MGB/VA | DAS Manual Curation |
| C0040610 | tramadol | CUI | MGB/VA | DAS Manual Curation |
| C0040808 | Treatment Protocols | CUI | MGB/VA | ONCE |
| C0041296 | Tuberculosis | CUI | MGB/VA | ONCE |
| C0041349 | Nephritis, Tubulointerstitial | CUI | MGB/VA | ONCE |
| C0041582 | Ulcer | CUI | MGB/VA | ONCE |
| C0041834 | Erythema | CUI | MGB/VA | ONCE |
| C0041976 | Urethritis | CUI | MGB/VA | ONCE |
| C0042014 | Urinalysis | CUI | MGB/VA | ONCE |
| C0042109 | Urticaria | CUI | MGB/VA | ONCE |
| C0042282 | Valgus deformity | CUI | MGB/VA | DAS Manual Curation |
| C0042384 | Vasculitis | CUI | MGB/VA | ONCE |
| C0042721 | Viral hepatitis | CUI | MGB/VA | ONCE |
| C0043047 | water | CUI | MGB/VA | ONCE |
| C0043251 | Wounds and Injuries | CUI | MGB/VA | DAS Manual Curation |
| C0043299 | Diagnostic radiologic examination | CUI | MGB/VA | ONCE |
| C0057476 | Depo-Medrol | CUI | MGB/VA | DAS Manual Curation |
| C0060926 | gabapentin | CUI | MGB/VA | DAS Manual Curation |
| C0063041 | leflunomide | CUI | MGB/VA | ONCE |
| C0079595 | Imaging Techniques | CUI | MGB/VA | ONCE |
| C0080078 | Range of Motion, Articular | CUI | MGB/VA | DAS Manual Curation |
| C0085435 | Arthritis, Reactive | CUI | MGB/VA | ONCE |
| C0085574 | Palindromic rheumatism | CUI | MGB/VA | DAS Manual Curation |
| C0085648 | Synovial Cyst | CUI | MGB/VA | DAS Manual Curation |
| C0085655 | Polymyositis | CUI | MGB/VA | ONCE |
| C0086437 | Joint laxity | CUI | MGB/VA | ONCE |
| C0086511 | Knee Replacement Arthroplasty (procedure) | CUI | MGB/VA | DAS Manual Curation |
| C0086565 | Liver Dysfunction | CUI | MGB/VA | ONCE |
| C0086787 | Percocet | CUI | MGB/VA | DAS Manual Curation |
| C0087111 | Therapeutic procedure | CUI | MGB/VA | DAS Manual Curation |
| C0087136 | Unmarried | CUI | MGB/VA | ONCE |
| C0117571 | Ferrlecit | CUI | MGB/VA | DAS Manual Curation |
| C0149756 | Fasciitis, Plantar | CUI | MGB/VA | DAS Manual Curation |
| C0149871 | Deep Vein Thrombosis | CUI | MGB/VA | ONCE |
| C0149910 | Intermittent joint effusion | CUI | MGB/VA | DAS Manual Curation |
| C0150220 | Range of motion exercise | CUI | MGB/VA | ONCE |
| C0150369 | Preventive monitoring | CUI | MGB/VA | DAS Manual Curation |
| C0151379 | Rheumatoid factor positive (finding) | CUI | MGB/VA | DAS Manual Curation |
| C0151434 | Biceps tendinitis | CUI | MGB/VA | DAS Manual Curation |
| C0151448 | Rotator Cuff Tendinitis | CUI | MGB/VA | DAS Manual Curation |
| C0151480 | Anti-nuclear factor positive | CUI | MGB/VA | ONCE |
| C0151735 | Injection Site Reaction | CUI | MGB/VA | ONCE |
| C0151766 | Liver function tests abnormal finding | CUI | MGB/VA | DAS Manual Curation |
| C0151937 | Rupture of tendon | CUI | MGB/VA | ONCE |
| C0152031 | Joint swelling | CUI | MGB/VA | ONCE |
| C0152054 | Therapeutic tactile stimulation | CUI | MGB/VA | DAS Manual Curation |
| C0152087 | Crystal Arthropathies | CUI | MGB/VA | ONCE |
| C0158026 | Monoarthritis | CUI | MGB/VA | ONCE |
| C0158328 | Trigger Finger Disorder | CUI | MGB/VA | DAS Manual Curation |
| C0162296 | Polyarthralgia | CUI | MGB/VA | ONCE |
| C0162298 | Joint stiffness | CUI | MGB/VA | ONCE |
| C0162323 | Polyarthritis | CUI | MGB/VA | ONCE |
| C0162429 | Malnutrition | CUI | MGB/VA | ONCE |
| C0163712 | Relate - vinyl resin | CUI | MGB/VA | DAS Manual Curation |
| C0168634 | BaseLine dental cement | CUI | MGB/VA | ONCE |
| C0175659 | Weight measurement scales | CUI | MGB/VA | DAS Manual Curation |
| C0175663 | Sounds device | CUI | MGB/VA | DAS Manual Curation |
| C0181904 | Monitor Device | CUI | MGB/VA | DAS Manual Curation |
| C0184301 | Vial device | CUI | MGB/VA | DAS Manual Curation |
| C0184511 | Improved | CUI | MGB/VA | DAS Manual Curation |
| C0184661 | Interventional procedure | CUI | MGB/VA | DAS Manual Curation |
| C0184958 | Toilet procedure | CUI | MGB/VA | DAS Manual Curation |
| C0185304 | Synovectomy | CUI | MGB/VA | ONCE |
| C0185317 | Arthroplasty, Replacement | CUI | MGB/VA | ONCE |
| C0187769 | Operative procedure on knee | CUI | MGB/VA | DAS Manual Curation |
| C0188413 | Operative procedure on foot | CUI | MGB/VA | DAS Manual Curation |
| C0199168 | Medical service | CUI | MGB/VA | DAS Manual Curation |
| C0200637 | Monocyte count procedure | CUI | MGB/VA | ONCE |
| C0201657 | C-reactive protein measurement | CUI | MGB/VA | ONCE |
| C0201660 | Rheumatoid Factor Measurement | CUI | MGB/VA | ONCE |
| C0201925 | Calcium measurement | CUI | MGB/VA | DAS Manual Curation |
| C0201975 | Creatinine measurement | CUI | MGB/VA | DAS Manual Curation |
| C0202202 | Protein measurement | CUI | MGB/VA | ONCE |
| C0202404 | Lidocaine measurement | CUI | MGB/VA | DAS Manual Curation |
| C0204854 | Arthrocentesis | CUI | MGB/VA | ONCE |
| C0205082 | Severe (severity modifier) | CUI | MGB/VA | DAS Manual Curation |
| C0205161 | Abnormal | CUI | MGB/VA | DAS Manual Curation |
| C0205400 | Thickened | CUI | MGB/VA | ONCE |
| C0206046 | Zofran | CUI | MGB/VA | DAS Manual Curation |
| C0206062 | Lung Diseases, Interstitial | CUI | MGB/VA | ONCE |
| C0220781 | Anabolism | CUI | MGB/VA | DAS Manual Curation |
| C0220787 | Endotracheal aspiration | CUI | MGB/VA | DAS Manual Curation |
| C0220929 | Mental tolerance | CUI | MGB/VA | DAS Manual Curation |
| C0221198 | Lesion | CUI | MGB/VA | DAS Manual Curation |
| C0221248 | Tophus | CUI | MGB/VA | ONCE |
| C0221423 | Illness (finding) | CUI | MGB/VA | ONCE |
| C0221785 | Pain in wrist | CUI | MGB/VA | DAS Manual Curation |
| C0231170 | Disability | CUI | MGB/VA | ONCE |
| C0231197 | Physiologic tolerance | CUI | MGB/VA | DAS Manual Curation |
| C0231218 | Malaise | CUI | MGB/VA | ONCE |
| C0231221 | Asymptomatic (finding) | CUI | MGB/VA | ONCE |
| C0231455 | Decreased flexion | CUI | MGB/VA | DAS Manual Curation |
| C0231456 | Abduction, function | CUI | MGB/VA | DAS Manual Curation |
| C0231528 | Myalgia | CUI | MGB/VA | ONCE |
| C0231589 | Limitation of joint movement | CUI | MGB/VA | DAS Manual Curation |
| C0231736 | Drawer sign | CUI | MGB/VA | DAS Manual Curation |
| C0231749 | Knee pain | CUI | MGB/VA | DAS Manual Curation |
| C0231790 | Thompson's test | CUI | MGB/VA | DAS Manual Curation |
| C0232117 | Pulse Rate | CUI | MGB/VA | DAS Manual Curation |
| C0233481 | Worried | CUI | MGB/VA | DAS Manual Curation |
| C0233494 | Tension | CUI | MGB/VA | DAS Manual Curation |
| C0234225 | Absence of pain | CUI | MGB/VA | DAS Manual Curation |
| C0234233 | Sore to touch | CUI | MGB/VA | ONCE |
| C0234238 | Ache | CUI | MGB/VA | ONCE |
| C0234255 | Night pain | CUI | MGB/VA | DAS Manual Curation |
| C0234856 | Speaking (activity) | CUI | MGB/VA | DAS Manual Curation |
| C0235108 | Feeling tense | CUI | MGB/VA | DAS Manual Curation |
| C0235439 | Ankle edema (finding) | CUI | MGB/VA | DAS Manual Curation |
| C0238551 | Left lower quadrant pain | CUI | MGB/VA | DAS Manual Curation |
| C0238656 | Ankle pain | CUI | MGB/VA | DAS Manual Curation |
| C0239134 | Productive Cough | CUI | MGB/VA | DAS Manual Curation |
| C0239833 | Hand pain | CUI | MGB/VA | ONCE |
| C0239946 | Fibrosis, Liver | CUI | MGB/VA | ONCE |
| C0240085 | JOINT ERYTHEMA | CUI | MGB/VA | DAS Manual Curation |
| C0240094 | Joint tenderness | CUI | MGB/VA | ONCE |
| C0240130 | Swollen knee region | CUI | MGB/VA | DAS Manual Curation |
| C0241040 | Pain of right shoulder joint | CUI | MGB/VA | DAS Manual Curation |
| C0241044 | SHOULDER TENDERNESS | CUI | MGB/VA | DAS Manual Curation |
| C0241760 | Wrist swelling | CUI | MGB/VA | DAS Manual Curation |
| C0242381 | Lyme Arthritis | CUI | MGB/VA | ONCE |
| C0242490 | Enthesopathy | CUI | MGB/VA | ONCE |
| C0242656 | Disease Progression | CUI | MGB/VA | ONCE |
| C0242708 | Antirheumatic Drugs, Disease-Modifying | CUI | MGB/VA | ONCE |
| C0242781 | disease transmission | CUI | MGB/VA | ONCE |
| C0245109 | anakinra | CUI | MGB/VA | DAS Manual Curation |
| C0253787 | acetyl 4-aminosalicylic acid | CUI | VA | DAS Manual Curation |
| C0262428 | Collagen-vascular disease | CUI | MGB/VA | ONCE |
| C0262926 | Medical History | CUI | MGB/VA | DAS Manual Curation |
| C0263591 | Drug-induced lupus erythematosus | CUI | MGB/VA | ONCE |
| C0263680 | Chronic arthritis | CUI | MGB/VA | ONCE |
| C0263907 | Capsulitis | CUI | MGB/VA | ONCE |
| C0263933 | Achilles tendinitis | CUI | MGB/VA | DAS Manual Curation |
| C0263962 | Olecranon bursitis | CUI | MGB/VA | ONCE |
| C0267374 | Idiopathic chronic inflammatory bowel disease | CUI | MGB/VA | DAS Manual Curation |
| C0275547 | Balanitis circinata | CUI | VA | DAS Manual Curation |
| C0277785 | Functional disorder | CUI | MGB/VA | DAS Manual Curation |
| C0277814 | Sitting position | CUI | MGB/VA | DAS Manual Curation |
| C0277964 | Subcutaneous crepitus | CUI | MGB/VA | DAS Manual Curation |
| C0278329 | Prescribed | CUI | MGB/VA | DAS Manual Curation |
| C0281481 | Anti-Tumor Necrosis Factor Therapy | CUI | MGB/VA | ONCE |
| C0281856 | Generalized aches and pains | CUI | MGB/VA | ONCE |
| C0302142 | Deformity | CUI | MGB/VA | DAS Manual Curation |
| C0308718 | CONTROL veterinary product | CUI | MGB/VA | DAS Manual Curation |
| C0309049 | favor | CUI | MGB/VA | DAS Manual Curation |
| C0311284 | Lipoid dermatoarthritis | CUI | VA | DAS Manual Curation |
| C0311392 | Physical findings | CUI | MGB/VA | DAS Manual Curation |
| C0332128 | Examined | CUI | MGB/VA | ONCE |
| C0332218 | Difficult (qualifier value) | CUI | MGB/VA | DAS Manual Curation |
| C0332219 | Easy | CUI | MGB/VA | DAS Manual Curation |
| C0332293 | Treated with | CUI | MGB/VA | ONCE |
| C0332447 | Morphologically abnormal structure (morphologic abnormality) | CUI | MGB/VA | ONCE |
| C0332448 | Infiltration | CUI | MGB/VA | ONCE |
| C0332461 | Plaque (lesion) | CUI | MGB/VA | DAS Manual Curation |
| C0332575 | Redness | CUI | MGB/VA | ONCE |
| C0332768 | Joint Subluxations | CUI | MGB/VA | ONCE |
| C0333068 | Flexion contracture | CUI | MGB/VA | ONCE |
| C0333243 | Pitting edema | CUI | MGB/VA | ONCE |
| C0333307 | Superficial ulcer | CUI | MGB/VA | ONCE |
| C0333361 | Acute inflammation | CUI | MGB/VA | DAS Manual Curation |
| C0333641 | Atrophic | CUI | MGB/VA | DAS Manual Curation |
| C0334094 | Proliferation (morphologic abnormality) | CUI | MGB/VA | DAS Manual Curation |
| C0343165 | Ankle joint effusion | CUI | MGB/VA | DAS Manual Curation |
| C0343166 | Knee joint effusion | CUI | MGB/VA | DAS Manual Curation |
| C0343167 | Wrist joint effusion | CUI | MGB/VA | DAS Manual Curation |
| C0344307 | Absence of pain sensation | CUI | MGB/VA | DAS Manual Curation |
| C0349707 | Aspiration-action | CUI | MGB/VA | DAS Manual Curation |
| C0358845 | Analgesics and non-steroidal anti-inflammatory drugs | CUI | MGB/VA | DAS Manual Curation |
| C0373527 | Acetaminophen Assay | CUI | MGB/VA | ONCE |
| C0376358 | Malignant neoplasm of prostate | CUI | MGB/VA | DAS Manual Curation |
| C0376569 | Fosamax | CUI | MGB/VA | DAS Manual Curation |
| C0376636 | Disease Management | CUI | MGB/VA | DAS Manual Curation |
| C0391850 | Physiologic pulse | CUI | MGB/VA | DAS Manual Curation |
| C0392197 | Physiologic warmth | CUI | MGB/VA | DAS Manual Curation |
| C0392367 | Physical contact | CUI | MGB/VA | DAS Manual Curation |
| C0392920 | Chemotherapy Regimen | CUI | MGB/VA | DAS Manual Curation |
| C0393022 | rituximab | CUI | MGB/VA | DAS Manual Curation |
| C0398650 | Immune thrombocytopenic purpura | CUI | MGB/VA | ONCE |
| C0405580 | Adrenal cortical hypofunction | CUI | MGB/VA | ONCE |
| C0406737 | Robinson nail dystrophy-deafness syndrome | CUI | MGB/VA | ONCE |
| C0409208 | Arthritis of hand | CUI | MGB/VA | ONCE |
| C0409210 | Arthritis of wrist | CUI | MGB/VA | ONCE |
| C0409338 | Flexion contracture - elbow | CUI | MGB/VA | DAS Manual Curation |
| C0409651 | Seropositive rheumatoid arthritis | CUI | MGB/VA | ONCE |
| C0409652 | Seronegative rheumatoid arthritis | CUI | MGB/VA | ONCE |
| C0409961 | Arthritis associated with another disorder | CUI | MGB/VA | DAS Manual Curation |
| C0409974 | Lupus Erythematosus | CUI | MGB/VA | ONCE |
| C0410000 | Overlap syndrome | CUI | MGB/VA | ONCE |
| C0410574 | Synovial Hypertrophy | CUI | MGB/VA | ONCE |
| C0413258 | Barotrauma of descent | CUI | MGB/VA | DAS Manual Curation |
| C0424577 | No complaints | CUI | MGB/VA | DAS Manual Curation |
| C0424578 | Psychological Well Being | CUI | MGB/VA | DAS Manual Curation |
| C0424653 | Weight symptom (finding) | CUI | MGB/VA | DAS Manual Curation |
| C0424755 | Fever symptoms (finding) | CUI | MGB/VA | ONCE |
| C0427008 | Stiffness | CUI | MGB/VA | ONCE |
| C0427244 | Joint deformity | CUI | MGB/VA | ONCE |
| C0427305 | Arthritis by pattern of joint involvement | CUI | MGB/VA | DAS Manual Curation |
| C0427896 | Crystal - human material | CUI | MGB/VA | DAS Manual Curation |
| C0439044 | Living Alone | CUI | MGB/VA | ONCE |
| C0439662 | Immune | CUI | MGB/VA | DAS Manual Curation |
| C0439775 | Elevation procedure | CUI | MGB/VA | DAS Manual Curation |
| C0441640 | Tapering - action | CUI | MGB/VA | DAS Manual Curation |
| C0441722 | Force | CUI | MGB/VA | DAS Manual Curation |
| C0441723 | Irritation | CUI | MGB/VA | ONCE |
| C0442735 | Nothing | CUI | MGB/VA | DAS Manual Curation |
| C0442739 | No status change | CUI | MGB/VA | ONCE |
| C0442743 | Noninflammatory | CUI | MGB/VA | ONCE |
| C0442797 | Decreasing | CUI | MGB/VA | DAS Manual Curation |
| C0442811 | Very low (qualifier value) | CUI | MGB/VA | DAS Manual Curation |
| C0442816 | Normal limits | CUI | MGB/VA | DAS Manual Curation |
| C0442874 | Neuropathy | CUI | MGB/VA | ONCE |
| C0442893 | Systemic disease | CUI | MGB/VA | ONCE |
| C0443146 | Autoimmune reaction | CUI | MGB/VA | ONCE |
| C0443343 | Unstable status | CUI | MGB/VA | DAS Manual Curation |
| C0444626 | Crystal Structure | CUI | MGB/VA | DAS Manual Curation |
| C0445223 | Related personal status | CUI | MGB/VA | DAS Manual Curation |
| C0445403 | Human patch material | CUI | MGB/VA | DAS Manual Curation |
| C0449752 | Ulnar deviation | CUI | MGB/VA | ONCE |
| C0449820 | Score | CUI | MGB/VA | DAS Manual Curation |
| C0450442 | Agent | CUI | MGB/VA | DAS Manual Curation |
| C0452240 | Physical therapy exercises | CUI | MGB/VA | DAS Manual Curation |
| C0454323 | Shoulder exercises | CUI | MGB/VA | DAS Manual Curation |
| C0455571 | H/O: rheumatoid arthritis | CUI | MGB/VA | DAS Manual Curation |
| C0455610 | H/O: surgery | CUI | MGB/VA | DAS Manual Curation |
| C0457086 | Morning stiffness - joint | CUI | MGB/VA | ONCE |
| C0460139 | Pressure (finding) | CUI | MGB/VA | DAS Manual Curation |
| C0475091 | Musculoskeletal system physical examination | CUI | MGB/VA | DAS Manual Curation |
| C0476273 | Respiratory distress | CUI | MGB/VA | ONCE |
| C0478530 | Examination and observation for unspecified reason | CUI | MGB/VA | DAS Manual Curation |
| C0483514 | Vicodin | CUI | MGB/VA | DAS Manual Curation |
| C0488564 | Review of systems:Finding:Point in time:^Patient:Narrative:Reported | CUI | MGB/VA | DAS Manual Curation |
| C0488565 | Review of systems:Finding:Point in time:^Patient:Nominal:Reported | CUI | MGB/VA | DAS Manual Curation |
| C0489534 | History of functional status | CUI | MGB/VA | DAS Manual Curation |
| C0489633 | Review of systems (procedure) | CUI | MGB/VA | DAS Manual Curation |
| C0494897 | Other rheumatoid arthritis | CUI | MGB/VA | ONCE |
| C0516979 | energy level | CUI | MGB/VA | DAS Manual Curation |
| C0517660 | Assessment of passive left elbow joint movement | CUI | MGB/VA | DAS Manual Curation |
| C0518015 | Hemoglobin measurement | CUI | MGB/VA | ONCE |
| C0518087 | Pain level | CUI | MGB/VA | DAS Manual Curation |
| C0518609 | Consideration | CUI | MGB/VA | DAS Manual Curation |
| C0518610 | Social warmth | CUI | MGB/VA | DAS Manual Curation |
| C0518656 | Chronic fatigue | CUI | MGB/VA | DAS Manual Curation |
| C0521144 | Seronegative | CUI | MGB/VA | ONCE |
| C0521516 | Polymyalgia | CUI | MGB/VA | DAS Manual Curation |
| C0523631 | Folic acid measurement | CUI | MGB/VA | ONCE |
| C0543419 | Sequela of disorder | CUI | MGB/VA | DAS Manual Curation |
| C0543467 | Operative Surgical Procedures | CUI | MGB/VA | DAS Manual Curation |
| C0543488 | Interested | CUI | MGB/VA | DAS Manual Curation |
| C0544452 | Disease remission | CUI | MGB/VA | DAS Manual Curation |
| C0548346 | Trauma assessment and care | CUI | MGB/VA | DAS Manual Curation |
| C0549433 | Surgical intervention (finding) | CUI | MGB/VA | DAS Manual Curation |
| C0553730 | Calcium pyrophosphate deposition disease | CUI | MGB/VA | ONCE |
| C0555903 | Total protein measurement | CUI | MGB/VA | ONCE |
| C0556895 | Combination electrotherapy | CUI | MGB/VA | DAS Manual Curation |
| C0557055 | Reassuring (procedure) | CUI | MGB/VA | DAS Manual Curation |
| C0557061 | Discussion (procedure) | CUI | MGB/VA | DAS Manual Curation |
| C0558145 | Skin appearance normal (finding) | CUI | MGB/VA | DAS Manual Curation |
| C0559169 | H/O: arthritis | CUI | MGB/VA | ONCE |
| C0560843 | Does lie down | CUI | MGB/VA | DAS Manual Curation |
| C0563625 | Agnosia for Pain | CUI | MGB/VA | ONCE |
| C0564405 | Feeling relief | CUI | MGB/VA | DAS Manual Curation |
| C0565683 | Able to carry | CUI | MGB/VA | DAS Manual Curation |
| C0567533 | Finding of lactation | CUI | MGB/VA | ONCE |
| C0574032 | Infusion procedures | CUI | MGB/VA | ONCE |
| C0574941 | Inflamed joint | CUI | MGB/VA | ONCE |
| C0574960 | Sacroiliitis | CUI | MGB/VA | ONCE |
| C0575545 | Shoulder joint - range of movement | CUI | MGB/VA | DAS Manual Curation |
| C0576091 | Deformity of knee joint | CUI | MGB/VA | DAS Manual Curation |
| C0576093 | Knee joint valgus deformity | CUI | MGB/VA | DAS Manual Curation |
| C0577559 | Mass of body structure | CUI | MGB/VA | ONCE |
| C0577599 | Swelling absent | CUI | MGB/VA | DAS Manual Curation |
| C0578014 | Hands normal | CUI | MGB/VA | DAS Manual Curation |
| C0581345 | Flare of rheumatoid arthritis | CUI | MGB/VA | ONCE |
| C0582103 | Medical Examination | CUI | MGB/VA | DAS Manual Curation |
| C0585962 | Seropositive erosive rheumatoid arthritis | CUI | MGB/VA | DAS Manual Curation |
| C0587240 | Erosion of bone | CUI | MGB/VA | ONCE |
| C0589120 | Follow-up status | CUI | MGB/VA | DAS Manual Curation |
| C0591159 | Benemid | CUI | MGB/VA | DAS Manual Curation |
| C0592278 | Zantac | CUI | MGB/VA | DAS Manual Curation |
| C0593507 | Advil | CUI | MGB/VA | DAS Manual Curation |
| C0595998 | Household composition | CUI | MGB/VA | ONCE |
| C0596764 | impression (attitude) | CUI | MGB/VA | DAS Manual Curation |
| C0596972 | Monitoring Device | CUI | MGB/VA | DAS Manual Curation |
| C0598463 | Functional Status | CUI | MGB/VA | ONCE |
| C0600117 | Does grip | CUI | MGB/VA | DAS Manual Curation |
| C0600139 | Prostate carcinoma | CUI | MGB/VA | DAS Manual Curation |
| C0600688 | Toxic effect | CUI | MGB/VA | DAS Manual Curation |
| C0611285 | APP protein, human | CUI | MGB/VA | DAS Manual Curation |
| C0666743 | infliximab | CUI | MGB/VA | ONCE |
| C0677042 | Pathology processes | CUI | MGB/VA | ONCE |
| C0677874 | In complete remission | CUI | MGB/VA | DAS Manual Curation |
| C0677932 | Progressive Neoplastic Disease | CUI | MGB/VA | ONCE |
| C0678176 | Neurontin | CUI | MGB/VA | DAS Manual Curation |
| C0678181 | Zocor | CUI | MGB/VA | DAS Manual Curation |
| C0679006 | Decision | CUI | MGB/VA | DAS Manual Curation |
| C0683525 | treatment options | CUI | MGB/VA | DAS Manual Curation |
| C0684239 | Emotional tenderness | CUI | MGB/VA | ONCE |
| C0684336 | Impaired health | CUI | MGB/VA | DAS Manual Curation |
| C0687702 | Cancer Remission | CUI | MGB/VA | DAS Manual Curation |
| C0699094 | Xylocaine | CUI | MGB/VA | DAS Manual Curation |
| C0699142 | Tylenol | CUI | MGB/VA | DAS Manual Curation |
| C0699177 | Plaquenil | CUI | MGB/VA | DAS Manual Curation |
| C0699187 | Valium | CUI | MGB/VA | DAS Manual Curation |
| C0699203 | Motrin | CUI | MGB/VA | DAS Manual Curation |
| C0699279 | Imuran | CUI | MGB/VA | DAS Manual Curation |
| C0699319 | Cytoxan | CUI | MGB/VA | DAS Manual Curation |
| C0699458 | Medrol | CUI | MGB/VA | DAS Manual Curation |
| C0699547 | Azulfidine | CUI | MGB/VA | DAS Manual Curation |
| C0699604 | Sandimmune | CUI | MGB/VA | DAS Manual Curation |
| C0699605 | Neoral | CUI | VA | DAS Manual Curation |
| C0699886 | Mechanical Treatments | CUI | MGB/VA | DAS Manual Curation |
| C0699958 | Voltaren | CUI | MGB/VA | DAS Manual Curation |
| C0700008 | Myochrysine | CUI | MGB/VA | DAS Manual Curation |
| C0700017 | Naprosyn | CUI | MGB/VA | DAS Manual Curation |
| C0700148 | Congestion | CUI | MGB/VA | ONCE |
| C0700198 | Pulmonary aspiration | CUI | MGB/VA | DAS Manual Curation |
| C0700594 | Radiculopathy | CUI | MGB/VA | DAS Manual Curation |
| C0700763 | Pravachol | CUI | MGB/VA | DAS Manual Curation |
| C0700777 | Prilosec | CUI | MGB/VA | DAS Manual Curation |
| C0701331 | Relafen | CUI | MGB/VA | DAS Manual Curation |
| C0702102 | Arthritis mutilans | CUI | VA | DAS Manual Curation |
| C0717758 | etanercept | CUI | MGB/VA | ONCE |
| C0718644 | Arava | CUI | MGB/VA | ONCE |
| C0719198 | Celebrex | CUI | MGB/VA | DAS Manual Curation |
| C0719517 | Correct brand of docusate-phenolphthalein | CUI | MGB/VA | DAS Manual Curation |
| C0719754 | Depen | CUI | VA | DAS Manual Curation |
| C0719949 | Diovan | CUI | MGB/VA | DAS Manual Curation |
| C0720099 | Duration brand of oxymetazoline | CUI | MGB/VA | DAS Manual Curation |
| C0720193 | Enbrel | CUI | MGB/VA | DAS Manual Curation |
| C0723011 | Relief brand of phenylephrine | CUI | MGB/VA | DAS Manual Curation |
| C0723012 | Remicade | CUI | MGB/VA | DAS Manual Curation |
| C0723362 | Sleep brand of diphenhydramine hydrochloride | CUI | MGB/VA | DAS Manual Curation |
| C0723460 | Stress bismuth subsalicylate | CUI | MGB/VA | DAS Manual Curation |
| C0723712 | Therapeutic brand of coal tar | CUI | MGB/VA | DAS Manual Curation |
| C0725066 | Advance -- medical device | CUI | MGB/VA | DAS Manual Curation |
| C0726642 | In Touch | CUI | MGB/VA | DAS Manual Curation |
| C0728786 | Solganal | CUI | MGB/VA | DAS Manual Curation |
| C0728827 | transfers | CUI | MGB/VA | DAS Manual Curation |
| C0728873 | Monitor brand of insecticide | CUI | MGB/VA | DAS Manual Curation |
| C0728976 | Control brand of phenylpropanolamine | CUI | MGB/VA | DAS Manual Curation |
| C0732355 | Rituxan | CUI | MGB/VA | DAS Manual Curation |
| C0740057 | Decadron | CUI | MGB/VA | DAS Manual Curation |
| C0740144 | Deltasone | CUI | MGB/VA | DAS Manual Curation |
| C0740335 | Wrist deformity | CUI | MGB/VA | DAS Manual Curation |
| C0740394 | Hyperuricemia | CUI | MGB/VA | ONCE |
| C0742906 | Elevated C-reactive protein | CUI | MGB/VA | ONCE |
| C0745538 | knee discomfort | CUI | MGB/VA | DAS Manual Curation |
| C0746617 | MONOARTICULAR ARTHRITIS | CUI | VA | DAS Manual Curation |
| C0746890 | new onset | CUI | MGB/VA | DAS Manual Curation |
| C0746919 | NO TREATMENT | CUI | MGB/VA | DAS Manual Curation |
| C0750394 | White blood cell count decreased | CUI | MGB/VA | DAS Manual Curation |
| C0750426 | White blood cell count increased (lab result) | CUI | MGB/VA | ONCE |
| C0751437 | Adenohypophyseal Diseases | CUI | MGB/VA | DAS Manual Curation |
| C0751438 | Posterior pituitary disease | CUI | MGB/VA | DAS Manual Curation |
| C0757844 | TNFSF13 protein, human | CUI | MGB/VA | DAS Manual Curation |
| C0795934 | Digitorenocerebral Syndrome | CUI | MGB/VA | ONCE |
| C0802632 | Complications:Type:Duration of the study:^Patient:Nominal | CUI | MGB/VA | DAS Manual Curation |
| C0813154 | Antibodies, in vitro diagnostic | CUI | MGB/VA | DAS Manual Curation |
| C0848332 | Spots on skin | CUI | MGB/VA | DAS Manual Curation |
| C0851162 | Infections of musculoskeletal system | CUI | MGB/VA | DAS Manual Curation |
| C0856432 | Shoulder injection | CUI | MGB/VA | DAS Manual Curation |
| C0860865 | Albumin normal | CUI | MGB/VA | DAS Manual Curation |
| C0871633 | desire | CUI | MGB/VA | DAS Manual Curation |
| C0872173 | Functional Disability | CUI | MGB/VA | DAS Manual Curation |
| C0876068 | Lidoderm | CUI | MGB/VA | DAS Manual Curation |
| C0877248 | Adverse event | CUI | MGB/VA | DAS Manual Curation |
| C0877521 | Chronic synovitis | CUI | MGB/VA | DAS Manual Curation |
| C0878195 | Abate | CUI | MGB/VA | DAS Manual Curation |
| C0879626 | Adverse effects | CUI | MGB/VA | DAS Manual Curation |
| C0885876 | X-rays, Homeopathic Preparations | CUI | MGB/VA | DAS Manual Curation |
| C0886296 | Nursing interventions | CUI | MGB/VA | DAS Manual Curation |
| C0919386 | Pathology procedure | CUI | MGB/VA | ONCE |
| C0935444 | centering | CUI | MGB/VA | DAS Manual Curation |
| C0939510 | Trexall | CUI | MGB/VA | DAS Manual Curation |
| C0949430 | Sulfazin | CUI | MGB/VA | DAS Manual Curation |
| C0949690 | Spondylarthritis | CUI | MGB/VA | ONCE |
| C0949691 | Spondylarthropathies | CUI | MGB/VA | ONCE |
| C0949766 | Physical therapy | CUI | MGB/VA | DAS Manual Curation |
| C0972401 | Boards (medical device) | CUI | MGB/VA | DAS Manual Curation |
| C0994475 | Pills | CUI | MGB/VA | DAS Manual Curation |
| C0994894 | Patch Dosage Form | CUI | MGB/VA | DAS Manual Curation |
| C1096593 | Shoulder discomfort | CUI | MGB/VA | DAS Manual Curation |
| C1114750 | Other medications:Finding:Point in time:^Patient:Nominal | CUI | MGB/VA | DAS Manual Curation |
| C1115771 | Other medications | CUI | MGB/VA | DAS Manual Curation |
| C1122087 | adalimumab | CUI | MGB/VA | ONCE |
| C1136179 | Hammer Toe | CUI | MGB/VA | DAS Manual Curation |
| C1142151 | Scleromalacia | CUI | MGB/VA | DAS Manual Curation |
| C1155266 | Inflammatory Response | CUI | MGB/VA | ONCE |
| C1170364 | Kineret | CUI | MGB/VA | DAS Manual Curation |
| C1171255 | Humira | CUI | MGB/VA | DAS Manual Curation |
| C1176468 | Erythrocyte sedimentation rate measurement | CUI | MGB/VA | ONCE |
| C1254351 | Pharmacologic Substance | CUI | MGB/VA | DAS Manual Curation |
| C1260969 | Ring device | CUI | MGB/VA | DAS Manual Curation |
| C1261287 | Stenosis | CUI | MGB/VA | ONCE |
| C1262148 | Grip strength decreased | CUI | MGB/VA | ONCE |
| C1262477 | Weight decreased | CUI | MGB/VA | ONCE |
| C1263855 | Lumbar radiculopathy | CUI | MGB/VA | DAS Manual Curation |
| C1265570 | Morphology within normal limits | CUI | MGB/VA | DAS Manual Curation |
| C1266765 | Tramadol measurement (procedure) | CUI | MGB/VA | DAS Manual Curation |
| C1268649 | New finding since previous mammogram | CUI | MGB/VA | DAS Manual Curation |
| C1271104 | Blood pressure finding | CUI | MGB/VA | DAS Manual Curation |
| C1272641 | Systemic arterial pressure | CUI | MGB/VA | DAS Manual Curation |
| C1272755 | Lowered (qualifier value) | CUI | MGB/VA | ONCE |
| C1272883 | Injection | CUI | MGB/VA | DAS Manual Curation |
| C1272892 | Intravenous infusion (product) | CUI | MGB/VA | DAS Manual Curation |
| C1273869 | Intervention regimes | CUI | MGB/VA | DAS Manual Curation |
| C1282310 | Intermittent pain | CUI | MGB/VA | DAS Manual Curation |
| C1287267 | Finding of platelet count | CUI | MGB/VA | DAS Manual Curation |
| C1290884 | Inflammatory disorder | CUI | MGB/VA | ONCE |
| C1291764 | Immune System Finding | CUI | MGB/VA | ONCE |
| C1293131 | Fusion procedure | CUI | MGB/VA | DAS Manual Curation |
| C1294065 | Macrophage count | CUI | MGB/VA | ONCE |
| C1298682 | Shoulder arthritis | CUI | MGB/VA | ONCE |
| C1299582 | Unable | CUI | MGB/VA | ONCE |
| C1299586 | Has difficulty doing (qualifier value) | CUI | MGB/VA | ONCE |
| C1301725 | Documented | CUI | MGB/VA | DAS Manual Curation |
| C1304888 | Pain control | CUI | MGB/VA | ONCE |
| C1305400 | Surgical patch | CUI | MGB/VA | DAS Manual Curation |
| C1306645 | Plain x-ray | CUI | MGB/VA | ONCE |
| C1315072 | Serology (antibodies and most antigens except blood bank and infectious agents) | CUI | MGB/VA | ONCE |
| C1321605 | Compliance behavior | CUI | MGB/VA | DAS Manual Curation |
| C1331418 | Comfort | CUI | MGB/VA | DAS Manual Curation |
| C1332206 | Adult Lymphoma | CUI | MGB/VA | ONCE |
| C1334928 | Necrotic changes (finding) | CUI | MGB/VA | ONCE |
| C1363945 | Therapy Object (animal model) | CUI | MGB/VA | DAS Manual Curation |
| C1373218 | Decreased Immunologic Activity [PE] | CUI | MGB/VA | ONCE |
| C1378554 | Crystals | CUI | MGB/VA | DAS Manual Curation |
| C1410088 | Still | CUI | MGB/VA | DAS Manual Curation |
| C1444648 | Offered | CUI | MGB/VA | DAS Manual Curation |
| C1444650 | Not needed | CUI | MGB/VA | DAS Manual Curation |
| C1444656 | Indicated | CUI | MGB/VA | ONCE |
| C1444783 | Instability | CUI | MGB/VA | DAS Manual Curation |
| C1446911 | Scheduling (procedure) | CUI | MGB/VA | DAS Manual Curation |
| C1448177 | TNF protein, human | CUI | MGB/VA | ONCE |
| C1453915 | cyclic citrullinated peptide | CUI | MGB/VA | ONCE |
| C1456454 | Amyloid Proteins | CUI | MGB/VA | DAS Manual Curation |
| C1456820 | Tumor Necrosis Factor-alpha | CUI | MGB/VA | ONCE |
| C1457868 | Worse | CUI | MGB/VA | DAS Manual Curation |
| C1457887 | Symptoms | CUI | MGB/VA | DAS Manual Curation |
| C1457907 | Surgical procedure finding | CUI | MGB/VA | ONCE |
| C1458156 | Recurrent Malignant Neoplasm | CUI | MGB/VA | DAS Manual Curation |
| C1509143 | Physical assessment findings | CUI | MGB/VA | DAS Manual Curation |
| C1513197 | Rheumatrex | CUI | MGB/VA | DAS Manual Curation |
| C1513374 | Moderate Adverse Event | CUI | MGB/VA | DAS Manual Curation |
| C1517205 | Flare | CUI | MGB/VA | DAS Manual Curation |
| C1518681 | Outcome of Therapy | CUI | MGB/VA | DAS Manual Curation |
| C1519275 | Grade 3 Severe Adverse Event | CUI | MGB/VA | DAS Manual Curation |
| C1522577 | follow-up | CUI | MGB/VA | DAS Manual Curation |
| C1522704 | Exercise Pain Management | CUI | MGB/VA | DAS Manual Curation |
| C1525443 | W flexion | CUI | MGB/VA | DAS Manual Curation |
| C1527305 | Feelings | CUI | MGB/VA | DAS Manual Curation |
| C1533128 | Body tissue patch material | CUI | MGB/VA | DAS Manual Curation |
| C1533157 | Block Specimens | CUI | MGB/VA | DAS Manual Curation |
| C1533685 | Injection procedure | CUI | MGB/VA | DAS Manual Curation |
| C1548484 | rheumatic fever vaccine | CUI | MGB/VA | ONCE |
| C1550457 | Normal Observation Interpretation | CUI | MGB/VA | DAS Manual Curation |
| C1550678 | Water Specimen | CUI | MGB/VA | DAS Manual Curation |
| C1551394 | normal Device Alert Level | CUI | MGB/VA | DAS Manual Curation |
| C1553188 | Hemolysis - observation | CUI | MGB/VA | ONCE |
| C1555457 | Provision of recurring care for chronic illness | CUI | MGB/VA | DAS Manual Curation |
| C1561560 | ambulatory encounter | CUI | MGB/VA | DAS Manual Curation |
| C1561581 | Allergy Severity - Severe | CUI | MGB/VA | DAS Manual Curation |
| C1561593 | Administrative Gender - Undifferentiated | CUI | MGB/VA | ONCE |
| C1565489 | Renal Insufficiency | CUI | MGB/VA | ONCE |
| C1565860 | PTGS2 protein, human | CUI | MGB/VA | DAS Manual Curation |
| C1609165 | tocilizumab | CUI | MGB/VA | DAS Manual Curation |
| C1609432 | manifestations of immunopathology | CUI | MGB/VA | ONCE |
| C1609538 | Latent Tuberculosis | CUI | MGB/VA | DAS Manual Curation |
| C1619634 | erythrocyte sedimentation rate result | CUI | MGB/VA | ONCE |
| C1619966 | abatacept | CUI | MGB/VA | DAS Manual Curation |
| C1657761 | local injection | CUI | MGB/VA | ONCE |
| C1692321 | Cellular infiltrate | CUI | MGB/VA | ONCE |
| C1692871 | Inflammatory polyarthritis | CUI | MGB/VA | ONCE |
| C1692886 | Arthritis, Bacterial | CUI | MGB/VA | ONCE |
| C1700021 | Orencia | CUI | MGB/VA | ONCE |
| C1704264 | Interleukin 1 Receptor Antagonist Protein | CUI | VA | DAS Manual Curation |
| C1704632 | Disease Response | CUI | MGB/VA | DAS Manual Curation |
| C1705245 | Coloring Excipient | CUI | MGB/VA | DAS Manual Curation |
| C1705365 | Dressing Dosage Form | CUI | MGB/VA | ONCE |
| C1705687 | Inversion Mutation Abnormality | CUI | MGB/VA | DAS Manual Curation |
| C1706085 | Block Dosage Form | CUI | MGB/VA | DAS Manual Curation |
| C1706307 | Irritation (finding) | CUI | MGB/VA | ONCE |
| C1706355 | Culture Dose Form | CUI | MGB/VA | DAS Manual Curation |
| C1707974 | Patch - Extended Release Film | CUI | MGB/VA | DAS Manual Curation |
| C1717255 | Edema:Finding:Point in time:^Patient:Ordinal | CUI | MGB/VA | DAS Manual Curation |
| C1718621 | W stress | CUI | MGB/VA | DAS Manual Curation |
| C1739148 | Acute synovitis | CUI | MGB/VA | DAS Manual Curation |
| C1744706 | intolerance to substance | CUI | MGB/VA | DAS Manual Curation |
| C1827449 | Drug Tapering | CUI | MGB/VA | DAS Manual Curation |
| C1844820 | Range of joint movement increased | CUI | MGB/VA | ONCE |
| C1846009 | Intrauterine growth restriction, metaphyseal dysplasia, adrenal hypoplasia congenita, and genital anomaly syndrome | CUI | MGB/VA | DAS Manual Curation |
| C1853193 | Recurrent skin infections | CUI | MGB/VA | DAS Manual Curation |
| C1856053 | Hydranencephaly with Renal Aplasia-Dysplasia | CUI | MGB/VA | DAS Manual Curation |
| C1859695 | Narrowed joint space | CUI | MGB/VA | ONCE |
| C1868955 | Synovitis wrist | CUI | MGB/VA | DAS Manual Curation |
| C1872109 | certolizumab pegol | CUI | MGB/VA | DAS Manual Curation |
| C1873497 | Normal assessment finding | CUI | MGB/VA | DAS Manual Curation |
| C1874451 | Basis | CUI | MGB/VA | DAS Manual Curation |
| C1883468 | Unstable Medical Device Problem | CUI | MGB/VA | DAS Manual Curation |
| C1947901 | Cancer Progression | CUI | MGB/VA | ONCE |
| C1947910 | Pulse phenomenon | CUI | MGB/VA | DAS Manual Curation |
| C1955473 | Others - Allergy | CUI | MGB/VA | DAS Manual Curation |
| C1956346 | Coronary Artery Disease | CUI | MGB/VA | ONCE |
| C1959609 | Erosion lesion | CUI | MGB/VA | ONCE |
| C1962945 | Radiographic imaging procedure | CUI | MGB/VA | ONCE |
| C1964257 | Observation - diagnostic procedure | CUI | MGB/VA | ONCE |
| C1979801 | Routine coag | CUI | MGB/VA | DAS Manual Curation |
| C2004436 | Activity therapy | CUI | MGB/VA | DAS Manual Curation |
| C2004491 | Cicatrix | CUI | MGB/VA | ONCE |
| C2016948 | soft tissue pain in foot | CUI | MGB/VA | DAS Manual Curation |
| C2025995 | cellulitis on exam (physical finding) | CUI | MGB/VA | ONCE |
| C2051415 | patient appears in no acute distress (physical finding) | CUI | MGB/VA | DAS Manual Curation |
| C2081614 | plan: problem (treatment) | CUI | MGB/VA | DAS Manual Curation |
| C2081627 | plan: surgery (treatment) | CUI | MGB/VA | DAS Manual Curation |
| C2091306 | doctor's orders: continue current therapy | CUI | MGB/VA | DAS Manual Curation |
| C2117111 | toe x-ray - deformity | CUI | MGB/VA | DAS Manual Curation |
| C2117331 | toe x-ray: deformity rotation | CUI | MGB/VA | DAS Manual Curation |
| C2127213 | thigh symptoms | CUI | MGB/VA | DAS Manual Curation |
| C2127484 | pain in lateral epicondyle of right humerus | CUI | MGB/VA | DAS Manual Curation |
| C2135970 | last seen [use onset date] | CUI | MGB/VA | DAS Manual Curation |
| C2142181 | Pain of left knee joint | CUI | MGB/VA | DAS Manual Curation |
| C2156946 | warmth of elbow | CUI | MGB/VA | DAS Manual Curation |
| C2167935 | Pain of left wrist | CUI | MGB/VA | DAS Manual Curation |
| C2186386 | reported history of knee replacement | CUI | MGB/VA | DAS Manual Curation |
| C2202235 | Pain of right knee joint | CUI | MGB/VA | DAS Manual Curation |
| C2221194 | multiple nodules | CUI | MGB/VA | DAS Manual Curation |
| C2228489 | examination of thyroid | CUI | MGB/VA | DAS Manual Curation |
| C2230397 | range of motion of wrist | CUI | MGB/VA | DAS Manual Curation |
| C2230421 | range of motion of elbow | CUI | MGB/VA | DAS Manual Curation |
| C2239122 | Social history of activities | CUI | MGB/VA | DAS Manual Curation |
| C2239184 | acquired hammer toe | CUI | MGB/VA | ONCE |
| C2242847 | activities of daily living (history) | CUI | MGB/VA | DAS Manual Curation |
| C2243116 | range of motion of shoulder | CUI | MGB/VA | DAS Manual Curation |
| C2266644 | subjective (symptom) | CUI | MGB/VA | DAS Manual Curation |
| C2316676 | Prosthetic arthroplasty of left hip | CUI | MGB/VA | DAS Manual Curation |
| C2317432 | Effusion (substance) | CUI | MGB/VA | DAS Manual Curation |
| C2343521 | Cimzia | CUI | MGB/VA | DAS Manual Curation |
| C2343929 | Tumor Necrosis Factor Blocker [EPC] | CUI | MGB/VA | DAS Manual Curation |
| C2347080 | Material Absorption | CUI | MGB/VA | DAS Manual Curation |
| C2347509 | Physical Shift | CUI | MGB/VA | ONCE |
| C2347761 | Childhood Myelodysplastic Syndrome | CUI | MGB/VA | ONCE |
| C2348252 | Diclofenac Sodium Gel | CUI | MGB/VA | DAS Manual Curation |
| C2350522 | Touch Perception | CUI | MGB/VA | DAS Manual Curation |
| C2353893 | golimumab | CUI | MGB/VA | DAS Manual Curation |
| C2364135 | Discomfort | CUI | MGB/VA | DAS Manual Curation |
| C2584297 | Sitting Function | CUI | MGB/VA | DAS Manual Curation |
| C2585021 | Referral to | CUI | MGB/VA | DAS Manual Curation |
| C2598165 | Grip Strength:-:Point in time:^Patient:- | CUI | MGB/VA | DAS Manual Curation |
| C2684024 | Simponi | CUI | MGB/VA | DAS Manual Curation |
| C2697310 | SARCOIDOSIS, SUSCEPTIBILITY TO, 1 (finding) | CUI | MGB/VA | ONCE |
| C2699477 | Crystal Present Or Absent (lab procedure) | CUI | MGB/VA | DAS Manual Curation |
| C2707028 | Functional status:-:Point in time:^Patient:- | CUI | MGB/VA | DAS Manual Curation |
| C2712134 | Actual Positive Comfort | CUI | MGB/VA | DAS Manual Curation |
| C2712334 | Actual Aspiration | CUI | MGB/VA | DAS Manual Curation |
| C2732281 | Secondary osteoarthritis | CUI | MGB/VA | DAS Manual Curation |
| C2740854 | Actemra | CUI | MGB/VA | DAS Manual Curation |
| C2825032 | Withdrawal (dysfunction) | CUI | MGB/VA | ONCE |
| C2825050 | Energy Absorption | CUI | MGB/VA | DAS Manual Curation |
| C2825055 | Recurrence (disease attribute) | CUI | MGB/VA | DAS Manual Curation |
| C2827071 | Unintentional Material Aspiration | CUI | MGB/VA | DAS Manual Curation |
| C2827483 | Fixed Specimen | CUI | MGB/VA | DAS Manual Curation |
| C2827774 | Current Therapy | CUI | MGB/VA | DAS Manual Curation |
| C2889385 | Rheumatoid factor positive rheumatoid arthritis | CUI | MGB/VA | DAS Manual Curation |
| C2911643 | Encounter due to family history of osteoporosis | CUI | MGB/VA | ONCE |
| C2911690 | Disease Controlled | CUI | MGB/VA | DAS Manual Curation |
| C2911692 | Response (communication) | CUI | MGB/VA | DAS Manual Curation |
| C2926602 | Discharge, body substance | CUI | MGB/VA | ONCE |
| C2926606 | Procedure findings:Finding:Point in time:^Patient:Narrative | CUI | MGB/VA | DAS Manual Curation |
| C2926735 | Duration | CUI | MGB/VA | DAS Manual Curation |
| C2930696 | tofacitinib | CUI | MGB/VA | DAS Manual Curation |
| C2936886 | Calcium [EPC] | CUI | MGB/VA | ONCE |
| C2939176 | feeling miserable | CUI | MGB/VA | DAS Manual Curation |
| C2945560 | Hemolysis (lab result) | CUI | MGB/VA | ONCE |
| C2945667 | Assessment of active left elbow joint movement | CUI | MGB/VA | DAS Manual Curation |
| C2947996 | Perform | CUI | MGB/VA | DAS Manual Curation |
| C2973287 | Clinical impression | CUI | MGB/VA | DAS Manual Curation |
| C2979880 | Subjective:Finding:Point in time:^Patient:Narrative | CUI | MGB/VA | DAS Manual Curation |
| C2984078 | A little bit | CUI | MGB/VA | DAS Manual Curation |
| C2984079 | Somewhat | CUI | MGB/VA | DAS Manual Curation |
| C2984081 | Very Much | CUI | MGB/VA | DAS Manual Curation |
| C2986546 | Target Lesion Identification | CUI | MGB/VA | DAS Manual Curation |
| C2987187 | Pleasant | CUI | MGB/VA | DAS Manual Curation |
| C3160715 | Mixed Cell Morphology | CUI | MGB/VA | DAS Manual Curation |
| C3173575 | Interpretation:Impression/interpretation of study:Point in time:^Patient:Nominal | CUI | MGB/VA | DAS Manual Curation |
| C3202977 | Analgesia | CUI | MGB/VA | DAS Manual Curation |
| C3203072 | Sulfazine | CUI | MGB/VA | DAS Manual Curation |
| C3242274 | new therapy | CUI | MGB/VA | DAS Manual Curation |
| C3244287 | School Grade | CUI | MGB/VA | DAS Manual Curation |
| C3244313 | Inactive Healthcare Coverage | CUI | MGB/VA | ONCE |
| C3263700 | Instructions:Text:Point in time:^Patient:Narrative | CUI | MGB/VA | DAS Manual Curation |
| C3263722 | Traumatic AND/OR non-traumatic injury | CUI | MGB/VA | ONCE |
| C3263723 | Traumatic injury | CUI | MGB/VA | DAS Manual Curation |
| C3272281 | American College of Cardiology/American Heart Association Lesion Complexity Score A | CUI | MGB/VA | ONCE |
| C3273461 | Remetinostat | CUI | MGB/VA | DAS Manual Curation |
| C3274571 | Clinical Study Follow-up | CUI | MGB/VA | DAS Manual Curation |
| C3279947 | NAIL DISORDER, NONSYNDROMIC CONGENITAL, 9 | CUI | MGB/VA | ONCE |
| C3463824 | MYELODYSPLASTIC SYNDROME | CUI | MGB/VA | ONCE |
| C3469186 | HEMOCHROMATOSIS, TYPE 1 | CUI | MGB/VA | ONCE |
| C3495458 | antibodies (medication) | CUI | MGB/VA | DAS Manual Curation |
| C3505045 | Xeljanz | CUI | MGB/VA | DAS Manual Curation |
| C3536731 | Massage Therapy | CUI | MGB/VA | ONCE |
| C3537192 | Tumor Necrosis Factor Inhibitors | CUI | MGB/VA | DAS Manual Curation |
| C3539106 | Sufficiently defined concept definition status (core metadata concept) | CUI | MGB/VA | ONCE |
| C3539125 | other medicated shampoos in ATC | CUI | MGB/VA | DAS Manual Curation |
| C3539781 | Progressive cGVHD | CUI | MGB/VA | ONCE |
| C3540032 | Vitamin IV solution additives | CUI | MGB/VA | ONCE |
| C3540037 | CALCIUM SUPPLEMENTS | CUI | MGB/VA | DAS Manual Curation |
| C3540038 | THYROID DIAGNOSTIC RADIOPHARMACEUTICALS | CUI | MGB/VA | DAS Manual Curation |
| C3540542 | Exacerbation of cGVHD | CUI | MGB/VA | ONCE |
| C3540676 | Blockade | CUI | MGB/VA | DAS Manual Curation |
| C3540704 | Antibiotics for systemic use | CUI | MGB/VA | DAS Manual Curation |
| C3540705 | Antifungal Antibiotics, Topical | CUI | MGB/VA | DAS Manual Curation |
| C3540706 | Antibiotic throat preparations | CUI | MGB/VA | DAS Manual Curation |
| C3540707 | Antibiotics FOR TREATMENT OF HEMORRHOIDS AND ANAL FISSURES FOR TOPICAL USE | CUI | MGB/VA | DAS Manual Curation |
| C3540708 | Antibiotics, Gynecological | CUI | MGB/VA | DAS Manual Curation |
| C3540709 | antibiotics, intestinal | CUI | MGB/VA | DAS Manual Curation |
| C3540710 | Antibiotics, ophthalmologic | CUI | MGB/VA | DAS Manual Curation |
| C3540772 | Enzymes FOR DISORDERS OF THE MUSCULO-SKELETAL SYSTEM | CUI | MGB/VA | ONCE |
| C3540840 | Sign or Symptom | CUI | MGB/VA | ONCE |
| C3541902 | Measured Tumor Identification | CUI | MGB/VA | ONCE |
| C3641827 | Agree | CUI | MGB/VA | DAS Manual Curation |
| C3651896 | Simponi Aria | CUI | MGB/VA | DAS Manual Curation |
| C3652465 | Interferon | CUI | MGB/VA | ONCE |
| C3662528 | Discogenic pain | CUI | MGB/VA | DAS Manual Curation |
| C3665472 | Chemotherapy | CUI | MGB/VA | ONCE |
| C3666917 | Fiore | CUI | VA | ONCE |
| C3668776 | Otrexup | CUI | MGB/VA | DAS Manual Curation |
| C3668946 | Activity (animal life circumstance) | CUI | MGB/VA | DAS Manual Curation |
| C3669037 | Rotation (malposition) (morphologic abnormality) | CUI | MGB/VA | ONCE |
| C3669138 | Visual inspection (procedure) | CUI | MGB/VA | DAS Manual Curation |
| C3687832 | Drugs - dental services | CUI | MGB/VA | DAS Manual Curation |
| C3714514 | Infection | CUI | MGB/VA | ONCE |
| C3714552 | Weakness | CUI | MGB/VA | ONCE |
| C3714578 | Fix | CUI | MGB/VA | ONCE |
| C3714611 | Calcium Drug Class | CUI | MGB/VA | DAS Manual Curation |
| C3714614 | Distention | CUI | MGB/VA | ONCE |
| C3714636 | Pneumonitis | CUI | MGB/VA | ONCE |
| C3714655 | On IV | CUI | MGB/VA | DAS Manual Curation |
| C3714660 | Trauma | CUI | MGB/VA | ONCE |
| C3806379 | Granulation of tissue | CUI | MGB/VA | ONCE |
| C3807158 | Limited flexion | CUI | MGB/VA | DAS Manual Curation |
| C3811652 | A Proliferation-Inducing Ligand Measurement | CUI | MGB/VA | DAS Manual Curation |
| C3811910 | combination - answer to question | CUI | MGB/VA | DAS Manual Curation |
| C3812891 | All of the Time | CUI | MGB/VA | ONCE |
| C3812897 | General medical service | CUI | MGB/VA | DAS Manual Curation |
| C3814778 | Hemolytic Index | CUI | MGB/VA | ONCE |
| C3829994 | Generally Satisfied or Pleased | CUI | MGB/VA | DAS Manual Curation |
| C3830105 | Flare-up | CUI | MGB/VA | ONCE |
| C3830544 | Delight | CUI | MGB/VA | DAS Manual Curation |
| C3833417 | Moderate activity | CUI | MGB/VA | ONCE |
| C3834263 | Inactive - answer to question | CUI | MGB/VA | ONCE |
| C3840669 | 2 to 3 times | CUI | MGB/VA | ONCE |
| C3840684 | Modification | CUI | MGB/VA | ONCE |
| C3840869 | Boggy | CUI | MGB/VA | DAS Manual Curation |
| C3840892 | Day 4 | CUI | MGB/VA | DAS Manual Curation |
| C3840932 | Other Reason | CUI | MGB/VA | DAS Manual Curation |
| C3841003 | No test | CUI | MGB/VA | ONCE |
| C3841448 | Much worse | CUI | MGB/VA | ONCE |
| C3841449 | Much better | CUI | MGB/VA | DAS Manual Curation |
| C3841647 | Slightly worse | CUI | MGB/VA | DAS Manual Curation |
| C3841788 | 5 or 6 | CUI | MGB/VA | DAS Manual Curation |
| C3841811 | Transplant | CUI | MGB/VA | ONCE |
| C3842330 | Manually | CUI | MGB/VA | ONCE |
| C3842396 | No difference | CUI | MGB/VA | ONCE |
| C3842480 | More than 50 | CUI | MGB/VA | ONCE |
| C3842639 | 1-2½ hours | CUI | MGB/VA | DAS Manual Curation |
| C3843066 | 2 to 3 days | CUI | MGB/VA | DAS Manual Curation |
| C3843615 | Come and go | CUI | MGB/VA | ONCE |
| C3843801 | 3 or 4 | CUI | MGB/VA | ONCE |
| C3844120 | Can Do With Some Difficulty | CUI | MGB/VA | DAS Manual Curation |
| C3844126 | Can Do Very Little | CUI | MGB/VA | DAS Manual Curation |
| C3844332 | Not sure | CUI | MGB/VA | DAS Manual Curation |
| C3844714 | No improvement | CUI | MGB/VA | ONCE |
| C3845941 | No limitation | CUI | MGB/VA | DAS Manual Curation |
| C3848678 | Rasuvo | CUI | MGB/VA | DAS Manual Curation |
| C3853089 | Moderately good | CUI | MGB/VA | DAS Manual Curation |
| C3853114 | Activity somewhat limited | CUI | MGB/VA | DAS Manual Curation |
| C3853527 | Complement (Immunology Procedures) | CUI | MGB/VA | ONCE |
| C3853727 | Tobacco user | CUI | MGB/VA | ONCE |
| C3854081 | Problem:Find:Pt:^Patient:Nom | CUI | MGB/VA | DAS Manual Curation |
| C3854333 | Narrowing | CUI | MGB/VA | ONCE |
| C3854544 | Pharyngalgia | CUI | MGB/VA | ONCE |
| C3864035 | Bilateral carpal tunnel syndrome | CUI | MGB/VA | ONCE |
| C3878897 | Reclining chair | CUI | MGB/VA | DAS Manual Curation |
| C3885145 | sarilumab | CUI | MGB/VA | DAS Manual Curation |
| C3887524 | Skin Erosion | CUI | MGB/VA | ONCE |
| C3887597 | Acquired trigger finger | CUI | MGB/VA | DAS Manual Curation |
| C3889124 | Only a Little | CUI | MGB/VA | ONCE |
| C3890554 | Physical Inactivity | CUI | MGB/VA | ONCE |
| C3891813 | Had No Pain | CUI | MGB/VA | DAS Manual Curation |
| C3897632 | Severity of Symptom Score 0 | CUI | MGB/VA | ONCE |
| C3899394 | Definitely | CUI | MGB/VA | DAS Manual Curation |
| C3900098 | Adult Myelodysplastic Syndrome | CUI | MGB/VA | ONCE |
| C4015731 | No erythema | CUI | MGB/VA | DAS Manual Curation |
| C4018905 | Too early | CUI | MGB/VA | ONCE |
| C4025363 | Subluxation of metacarpal phalangeal joints | CUI | MGB/VA | DAS Manual Curation |
| C4034144 | Methotrexate Injection | CUI | MGB/VA | DAS Manual Curation |
| C4035626 | 3 times | CUI | MGB/VA | ONCE |
| C4035627 | 2 times | CUI | MGB/VA | ONCE |
| C4036115 | Very mild | CUI | MGB/VA | DAS Manual Curation |
| C4036205 | Ambulate | CUI | MGB/VA | DAS Manual Curation |
| C4044947 | baricitinib | CUI | MGB/VA | DAS Manual Curation |
| C4047978 | Rituximab therapy | CUI | MGB/VA | ONCE |
| C4048292 | Immobilization test | CUI | MGB/VA | ONCE |
| C4048329 | Immunosuppression | CUI | MGB/VA | ONCE |
| C4049139 | A Little Better | CUI | MGB/VA | DAS Manual Curation |
| C4049938 | Physical Activity Measurement | CUI | MGB/VA | DAS Manual Curation |
| C4050465 | Severe Extremity Pain | CUI | MGB/VA | ONCE |
| C4055223 | Clinical Response | CUI | MGB/VA | DAS Manual Curation |
| C4062321 | Functional impairment | CUI | MGB/VA | ONCE |
| C4068834 | Shiny | CUI | MGB/VA | ONCE |
| C4082130 | Prepared (qualifier value) | CUI | MGB/VA | ONCE |
| C4084203 | Improved - answer to question | CUI | MGB/VA | ONCE |
| C4084772 | Complement dependent cytotoxicity | CUI | MGB/VA | ONCE |
| C4085643 | Moderate Response | CUI | MGB/VA | ONCE |
| C4086268 | Exacerbation | CUI | MGB/VA | DAS Manual Curation |
| C4238109 | Inflectra | CUI | MGB/VA | DAS Manual Curation |
| C4264481 | 4 times | CUI | MGB/VA | ONCE |
| C4279925 | Total Joint Replacement | CUI | MGB/VA | ONCE |
| C4285457 | Fibrosis Assessment | CUI | MGB/VA | ONCE |
| C4285959 | Erosive arthritis | CUI | MGB/VA | ONCE |
| C4290029 | A Fair Amount | CUI | MGB/VA | DAS Manual Curation |
| C4316895 | Anaphylactic shock | CUI | MGB/VA | ONCE |
| C4318616 | Usually Need Help from Another Person for Arising | CUI | MGB/VA | ONCE |
| C4476350 | Kevzara | CUI | MGB/VA | DAS Manual Curation |
| C4476688 | Aggravated by activity | CUI | MGB/VA | DAS Manual Curation |
| C4484264 | Clark | CUI | MGB/VA | ONCE |
| C4490037 | Renflexis | CUI | VA | DAS Manual Curation |
| C4521229 | Neck Pain Score 6 | CUI | MGB/VA | ONCE |
| C4521576 | Neck Pain Score 0 | CUI | MGB/VA | DAS Manual Curation |
| C4522323 | Designated Blood Product Donation | CUI | MGB/VA | ONCE |
| C4534505 | Both reactive | CUI | VA | DAS Manual Curation |
| C4551516 | Hip pain | CUI | MGB/VA | DAS Manual Curation |
| C4551529 | Renal Dialysis | CUI | MGB/VA | ONCE |
| C4551577 | Crystal - natural material | CUI | MGB/VA | DAS Manual Curation |
| C4551889 | Creatinine metabolic function | CUI | MGB/VA | DAS Manual Curation |
| C4552279 | Tissue injury | CUI | MGB/VA | ONCE |
| C4552766 | Miscarriage | CUI | MGB/VA | ONCE |
| C4553026 | Calcium metabolic function | CUI | MGB/VA | DAS Manual Curation |
| C4553821 | Feels | CUI | MGB/VA | DAS Manual Curation |
| C4553887 | Biopharmaceuticals | CUI | MGB/VA | DAS Manual Curation |
| C4554465 | examination of kidney | CUI | MGB/VA | ONCE |
| C4554794 | Swollen joint:Anatomy:Point in time:^Patient:Nominal | CUI | MGB/VA | DAS Manual Curation |
| C4694844 | Olumiant | CUI | MGB/VA | DAS Manual Curation |
| C4698491 | Rather | CUI | MGB/VA | DAS Manual Curation |
| C4699158 | Increased risk | CUI | MGB/VA | ONCE |
| C4699190 | A pattern | CUI | MGB/VA | ONCE |
| C4706353 | DAS - Disease Activity Score | CUI | MGB/VA | DAS Manual Curation |
| C4716882 | Course of treatment | CUI | MGB/VA | ONCE |
| C4721453 | Peripheral Nervous System Diseases | CUI | MGB/VA | ONCE |
| C4721532 | Lymphoma, Non-Hodgkin, Familial | CUI | MGB/VA | ONCE |
| C4721829 | Radiographic Examination | CUI | MGB/VA | ONCE |
| C4722257 | STEPS to Enhance Physical Activity | CUI | MGB/VA | DAS Manual Curation |
| C4722398 | Patch Dosage Form Category | CUI | MGB/VA | DAS Manual Curation |
| C4722408 | Reactive Therapy | CUI | MGB/VA | ONCE |
| C4722602 | Underlying | CUI | MGB/VA | DAS Manual Curation |
| C4723839 | irPD (Immune-Related Response Criteria) | CUI | MGB/VA | ONCE |
| C4724253 | subharmonic aided pressure estimation | CUI | MGB/VA | DAS Manual Curation |
| C4726929 | upadacitinib | CUI | MGB/VA | DAS Manual Curation |
| C4738113 | Fatalities | CUI | MGB/VA | ONCE |
| C4740651 | Near remission | CUI | MGB/VA | DAS Manual Curation |
| C4740690 | Mild disease | CUI | MGB/VA | ONCE |
| C4740691 | Moderate disease | CUI | MGB/VA | ONCE |
| C4740692 | Severe disease | CUI | MGB/VA | ONCE |
| C4759845 | CTCAE v4 Grade 2 | CUI | MGB/VA | DAS Manual Curation |
| C4760706 | Increased circulating band cell count | CUI | MGB/VA | ONCE |
| C4763675 | Single Agent Therapy | CUI | MGB/VA | ONCE |
| C4763962 | Shoulder Replacement Surgery | CUI | MGB/VA | DAS Manual Curation |
| C5141269 | Increased pain | CUI | MGB/VA | DAS Manual Curation |
| C5196212 | Rinvoq | CUI | MGB/VA | DAS Manual Curation |
| C5200994 | Increased circulating prolactin concentration | CUI | MGB/VA | ONCE |
| C5201148 | Moderate | CUI | MGB/VA | ONCE |
| C5201282 | Adalimumab (drug assay) | CUI | MGB/VA | DAS Manual Curation |
| C5201962 | Infliximab (drug assay) | CUI | MGB/VA | DAS Manual Curation |
| C5202796 | Intensity and Distress 1 | CUI | MGB/VA | DAS Manual Curation |
| C5202916 | IPSS-R Risk Category Very High | CUI | MGB/VA | ONCE |
| C5202917 | IPSS-R Risk Category Very Low | CUI | MGB/VA | DAS Manual Curation |
| C5202921 | RECIL PD | CUI | MGB/VA | ONCE |
| C5202924 | Global Progressive Disease in Skin | CUI | MGB/VA | ONCE |
| C5202951 | Very Well | CUI | MGB/VA | DAS Manual Curation |
| C5202952 | Describes Very Well | CUI | MGB/VA | DAS Manual Curation |
| C5202953 | Can Do Very Well | CUI | MGB/VA | DAS Manual Curation |
| C5202991 | IMWG Progressive Disease | CUI | MGB/VA | ONCE |
| C5203119 | Intensity and Distress 5 | CUI | MGB/VA | DAS Manual Curation |
| C5203232 | Monoclonal Antibody [EPC] | CUI | MGB/VA | ONCE |
| C5203527 | Methotrexate Liquid Injection | CUI | MGB/VA | DAS Manual Curation |
| C5236002 | Increased (finding) | CUI | MGB/VA | DAS Manual Curation |
| C5236984 | Responsive Disease | CUI | MGB/VA | DAS Manual Curation |
| C5241184 | Intraarticular (intended site) | CUI | MGB/VA | ONCE |
| C5394288 | No rash | CUI | MGB/VA | DAS Manual Curation |
| C5395497 | Normal neck region | CUI | MGB/VA | DAS Manual Curation |
| C5399953 | Methotrexate Drug Assay | CUI | MGB/VA | DAS Manual Curation |
| C5401219 | Specified Cell Count Test | CUI | MGB/VA | DAS Manual Curation |
| C5401414 | Leflunomide drug assay | CUI | MGB/VA | DAS Manual Curation |
| C5420783 | Joint Involvement | CUI | MGB/VA | ONCE |
| C5425799 | All other | CUI | MGB/VA | DAS Manual Curation |
| C5441521 | Complaint (finding) | CUI | MGB/VA | ONCE |
| C5441745 | Abnormal pulmonary interstitial morphology | CUI | MGB/VA | ONCE |
| C5442019 | Pain of left knee region | CUI | MGB/VA | DAS Manual Curation |
| C5442020 | Pain of right knee region | CUI | MGB/VA | DAS Manual Curation |
| C5452990 | Helped | CUI | MGB/VA | DAS Manual Curation |
| C5545231 | Pool Therapy | CUI | MGB/VA | DAS Manual Curation |
| C1855652 | Fetus Small for Gestational Age | CUI | MGB | ONCE |
| C4704669 | Amjevita | CUI | MGB | DAS Manual Curation |
| C4704670 | Cyltezo | CUI | MGB | DAS Manual Curation |
