## Supplemental Figures for "Inferring rheumatoid arthritis disease activity status from the electronic health records across health systems to enable real-world data studies"

**Supplemental Figure 1.** Top 30 features with highest importance at the top among 263 features used in the algorithm developed based on MGB EHR data trained with BRASS DAS28-CRP data.

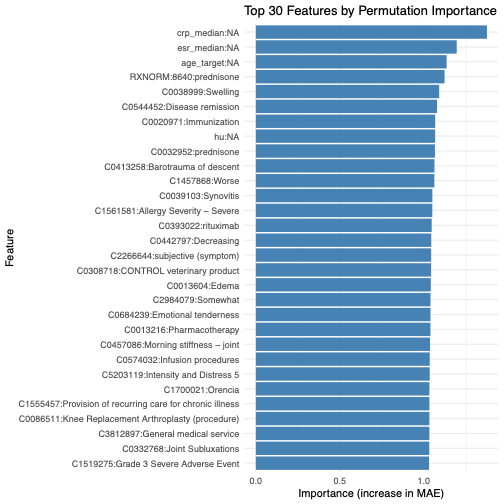

**Supplemental Figure 2.** Top 30 features with highest importance at the top among 518 features used in the algorithm developed with VA EHR data trained with VARA DAS28-CRP data.

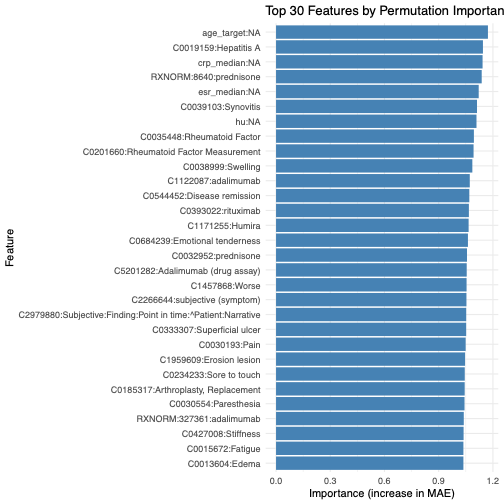

**Supplemental Figure 3.** Top 30 features with highest importance at the top among 211 used in the algorithm developed with MGB EHR data trained with BRASS DAS28-CRP data using only common features available in the MGB and VA EHRs.

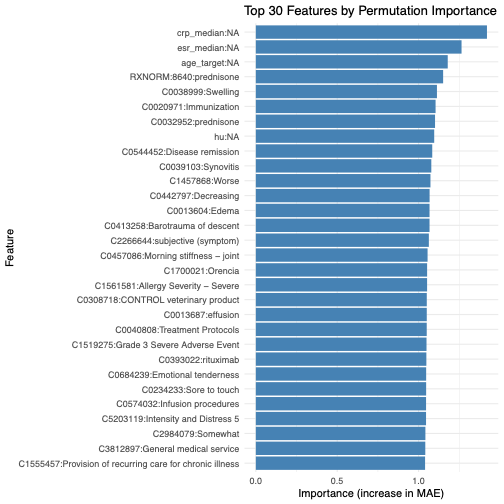

**Supplemental Figure 4.** Top 30 features with the highest importance at the top among 211 used in the algorithm developed with VA EHR data trained with VARA DAS28-CRP data using only common features available in MGB and VA EHRs.

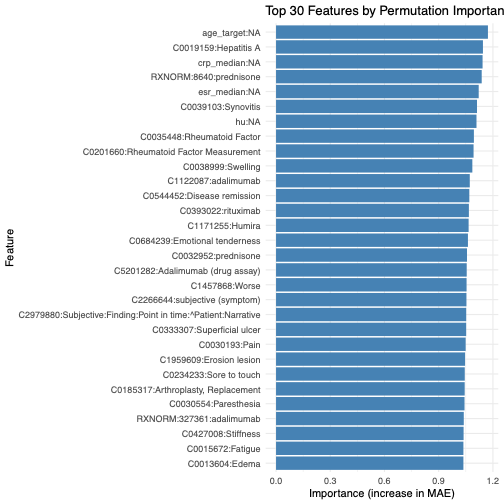
