## Supplemental Table 3 for "Inferring rheumatoid arthritis disease activity status from the electronic health records across health systems to enable real-world data studies"

**Supplemental Table 3.** Codified MACE definition.

|  | **ICD-9 Diagnosis** | **ICD-10 Diagnosis** |  |  |
| --- | --- | --- | --- | --- |
| *Diagnosis* |  |  |  |  |
| Myocardial infarction (MI) | 410.% | I21.0%, I21.1%, I21.2%, I21.29, I21.3, I21.4, I21.9, I21.A%, I21.B |  |  |
| Ischemic stroke | 433.%, 434.%, 435.%, 436.%, 437.1%, 437.9% | I65.1, I66.09, I66.19, I66.29, I67.81, I67.82, I67.848, I67.89, I67.9, G45.0, G45.1, G45.8, G45.9 |  |  |
|  | **ICD-9 Procedure** | **ICD-10 Procedure** | **CPT** | **DRG** |
| *Procedures* |  |  |  |  |
| Coronary artery bypass graft (CABG) | 36.1%, 36.2% | 0210093, 02100A3, 02100J3, 02100K3, 02100Z3, 0210493, 02104A3, 02104J3, 02104K3, 02104Z3, 021K0Z8, 021K0Z9, 021K0ZC, 021K0ZW, 021K4Z8, 021K4Z9, 021K4ZC, 021K4ZW, 021L0Z8, 021L0Z9, 021L0ZC, 021L4Z8, 021L4Z9, 021L4ZC | 33510-33536, 33545, 33572 | 106, 107, 109, 547, 548, 549, 550 |
| Percutaneous coronary intervention (PCI) | 00.66 | 02703ZZ, 02704ZZ, 02713ZZ, 02714ZZ, 02723ZZ , 02724ZZ, 02733ZZ, 02734ZZ | 92980-92982, 92995, 92997, 92982-92984 |  |
| Percutaenous transluminal coronary angioplasty (PTCA) | 36.03 | 02700ZZ, 02710ZZ, 02720ZZ, 02730ZZ, 02C00ZZ, 02C10ZZ, 02C20ZZ, 02C30ZZ |  |  |
| Stent | 00.66, 36.03, 36.06, 36.07, 36.09 | 02700D6, 02700DZ, 02700T6, 02700TZ, 02703D6, 02703DZ, 02703T6, 02703TZ, 02704D6, 02704DZ, 02704T6, 02704TZ, 02710D6, 02710DZ, 02710T6, 02710TZ, 02713D6, 02713DZ, 02713T6, 02713TZ, 02714D6, 02714DZ, 02714T6, 02714TZ, 02720D6, 02720DZ, 02720T6, 02720TZ, 02723D6, 02723DZ, 02723T6, 02723TZ, 02724D6, 02724DZ, 02724T6, 02724TZ, 02730D6, 02730DZ, 02730T6, 02730TZ, 02733D6, 02733DZ, 02733T6, 02733TZ, 02734D6, 02734DZ, 02734T6, 02734TZ,0270046, 027004Z, 0270346, 027034Z, 0270446, 027044Z, 0271046, 027104Z, 0271346, 027134Z, 0271446, 027144Z, 0272046, 027204Z, 0272346, 027234Z, 0272446, 027244Z, 0273046, 027304Z, 0273346, 027334Z, 0273446, 027344Z, 02C03ZZ, 02C04ZZ, 02C13ZZ, 02C14ZZ, 02C23ZZ, 02C24ZZ, 02C33ZZ, 02C34ZZ, 02703ZZ |  | 112, 555, 516, 517, 518, 556, 557, 558 |
